# Recessive variants in the intergenic *NOS1AP-C1orf226* locus cause monogenic kidney disease responsive to anti-proteinuric treatment

**DOI:** 10.1101/2024.03.17.24303374

**Authors:** Florian Buerger, Daanya Salmanullah, Lorrin Liang, Victoria Gauntner, Kavita Krueger, Maggie Qi, Vineeta Sharma, Alexander Rubin, David Ball, Katharina Lemberg, Ken Saida, Lea Maria Merz, Sanja Sever, Biju Issac, Liang Sun, Sergio Guerrero-Castillo, Nephrotic Syndrome Study Network (NEPTUNE), Alexis C. Gomez, Michelle T. McNulty, Matthew G. Sampson, Mohamed H. Al-Hamed, Mohammed M. Saleh, Mohamed Shalaby, Jameela Kari, James P. Fawcett, Friedhelm Hildebrandt, Amar J. Majmundar

## Abstract

In genetic disease, an accurate expression landscape of disease genes and faithful animal models will enable precise genetic diagnoses and therapeutic discoveries, respectively. We previously discovered that variants in *NOS1AP*, encoding nitric oxide synthase 1 (NOS1) adaptor protein, cause monogenic nephrotic syndrome (NS). Here, we determined that an intergenic splice product of N*OS1AP*/*Nos1ap* and neighboring *C1orf226/Gm7694*, which precludes NOS1 binding, is the predominant isoform in mammalian kidney transcriptional and proteomic data. *Gm7694^-/-^* mice, whose allele exclusively disrupts the intergenic product, developed NS phenotypes. In two human NS subjects, we identified causative *NOS1AP* splice variants, including one predicted to abrogate intergenic splicing but initially misclassified as benign based on the canonical transcript. Finally, by modifying genetic background, we generated a faithful mouse model of *NOS1AP*-associated NS, which responded to anti-proteinuric treatment. This study highlights the importance of intergenic splicing and a potential treatment avenue in a mendelian disorder.

## INTRODUCTION

Nephrotic syndrome (NS) is a leading cause of chronic kidney disease in children (*1*). NS presents with edema, hypoalbuminemia, and proteinuria, arising from disruption of the glomerular filtration barrier comprised primarily of interconnected podocytes (*2*). Treatment resistant NS (steroid-resistant NS or SRNS) frequently progresses to end-stage kidney disease concomitant with podocyte loss (*1–3*). Mendelian genetic forms represent the most severe subset of NS (*4–7*).

Monogenic NS genes are predominantly expressed in glomerular podocytes (*8*), encoding proteins essential for podocyte development or homeostasis (*2*, *9*). Patient variants in disease proteins impair podocyte structure and function (*2*, *8*), causing podocytopathies. For example, variants in >10 genes encoding actin cytoskeleton regulators have been shown to cause nephrotic syndrome in humans and mice (*10–19*). Despite the interconnected nature of these genetic etiologies, specific treatments are lacking.

Causative genetic variants are detected in 11-30% of SRNS cases (*4–7*). For families lacking a genetic diagnosis, variant classification can be hindered by transcript annotation based on non-kidney tissues. Understanding kidney-specific isoform expression will facilitate discovery of novel disease variants and genes, broadening our mechanistic understanding of NS and impacting clinical care for kidney disease patients with an established genetic diagnosis.

We, previously, discovered that recessive variants in *NOS1AP/Nos1ap*, affecting N-terminal domains, caused early onset NS in human subjects and a podocytopathy in C57BL/6-*Nos1ap^Ex3-/Ex3-^* mice (**Figure S1**) (*20*). This locus encodes nitric oxide synthase 1 adaptor protein with an N-terminal phosphotyrosine binding (PTB) domain, and C-terminal PDZ binding domain (PDZ-BD), through which it interacts with NOS1 (**Figure S1**) (*21–26*). We determined that NOS1AP co-localized with actin-based filopodia in podocytes and with podocyte marker nephrin in kidney sections. NS patient variants impaired filopodia formation and podocyte migration (*20*). NOS1AP acted upstream of the NS-associated CDC42/formin pathway, supporting its key role in the actin regulatory network (*20*).

While the C-terminal NOS1AP-NOS1 interaction is important for neuronal NO signaling and required for hippocampal neuron-mediated anxiolysis (*27*, *28*), impairing NOS1 function did not disrupt NOS1AP-dependent actin remodeling in podocytes in our study (*20*), suggesting an NOS1-independent role for NOS1AP in NS.

We previously described an additional intergenic *Nos1ap* transcript in rat tissues generated from splicing between the 5’ region of canonical exon 10 and two coding exons of neighboring open reading frame *LOC100361087* (*C1orf226* in humans and *Gm7694* in mice) that was most highly expressed in olfactory bulb tissue (*23*, *24*, *26*). This alternative transcript encodes a distinct C-terminal domain lacking the PDZ-BD that mediates NOS1 interactions (*23–26*). The role of this alternative transcript in kidney physiology and disease is unknown.

In the current study, we delineate an intergenic splice isoform of N*OS1AP*/*Nos1ap* and neighboring locus *C1orf226/Gm7694* by interrogation of transcriptional and proteomic data from human and murine kidneys. The intergenic form predominates over the canonical transcript in adult kidney tissue. We, next, hypothesized that novel variants in *NOS1AP/Nos1ap* cause NS and expand the genotype-phenotype relationship between this locus and kidney disease. *Gm7694^-/-^* mice, predicted to exclusively disrupt the intergenic *NOS1AP* splice product, developed a podocytopathy. These findings were mirrored in novel *Nos1ap^Ex4-/Ex4-^* mice with an early out-of-frame deletion and consistent with our published *Nos1ap^Ex3-/Ex3-^* in-frame deletion mouse model. Supporting the role of the NOS1AP C-terminus in humans, we identified two novel recessive variants in *NOS1AP*, predicted to impair splicing of the penultimate coding exon and the intergenic product, in two cases of pediatric onset NS. Collectively, this suggests that novel variants, which impact novel regions and splice isoforms of *NOS1AP/Nos1ap*, lead to kidney disease. To evaluate the impact of genetic modifying factors, the *Nos1ap^Ex3-^* allele was bred from a C57BL/6 background onto the FVB/N background. FVB/N-*Nos1ap^Ex3-/Ex3-^* mice exhibited 10-fold higher albuminuria than C57BL/6-*Nos1ap^Ex3-/Ex3-^* mice and developed kidney dysfunction, supporting the relevance of genetic background for *Nos1ap*-variant associated disease. Finally, FVB/N-*Nos1ap^Ex3-/Ex3-^* mice were treated with Renin-Angiotensin-Aldosterone-System (RAAS) inhibitor lisinopril, which reduced albuminuria and prevented premature death. Overall, our findings demonstrate that variants in the intergenic *NOS1AP-C1orf226* locus cause a podocytopathy responsive to pharmacologic treatment.

## RESULTS

### Intergenic splice product of *NOS1AP/Nos1ap* in human and mouse kidney

We, previously, described an intergenic splice product of the rat ortholog *Nos1ap* across multiple tissues with highest levels in olfactory bulb tissue and low expression in brain cortex and hippocampus (*23*, *24*, *26*). This alternative transcript encodes a distinct C-terminal domain lacking the PDZ-BD that mediates NOS1 interactions (*23–26*) (**Figure 1A, S1**). We sought to study this splice isoform further in mammalian kidneys, as (i) its product may mediate distinct protein interaction partners and biological roles than the canonical protein, and (ii) interpretation of genetic variants in individuals could be dramatically altered in light of tissue-specific isoforms.

**Figure 1.**
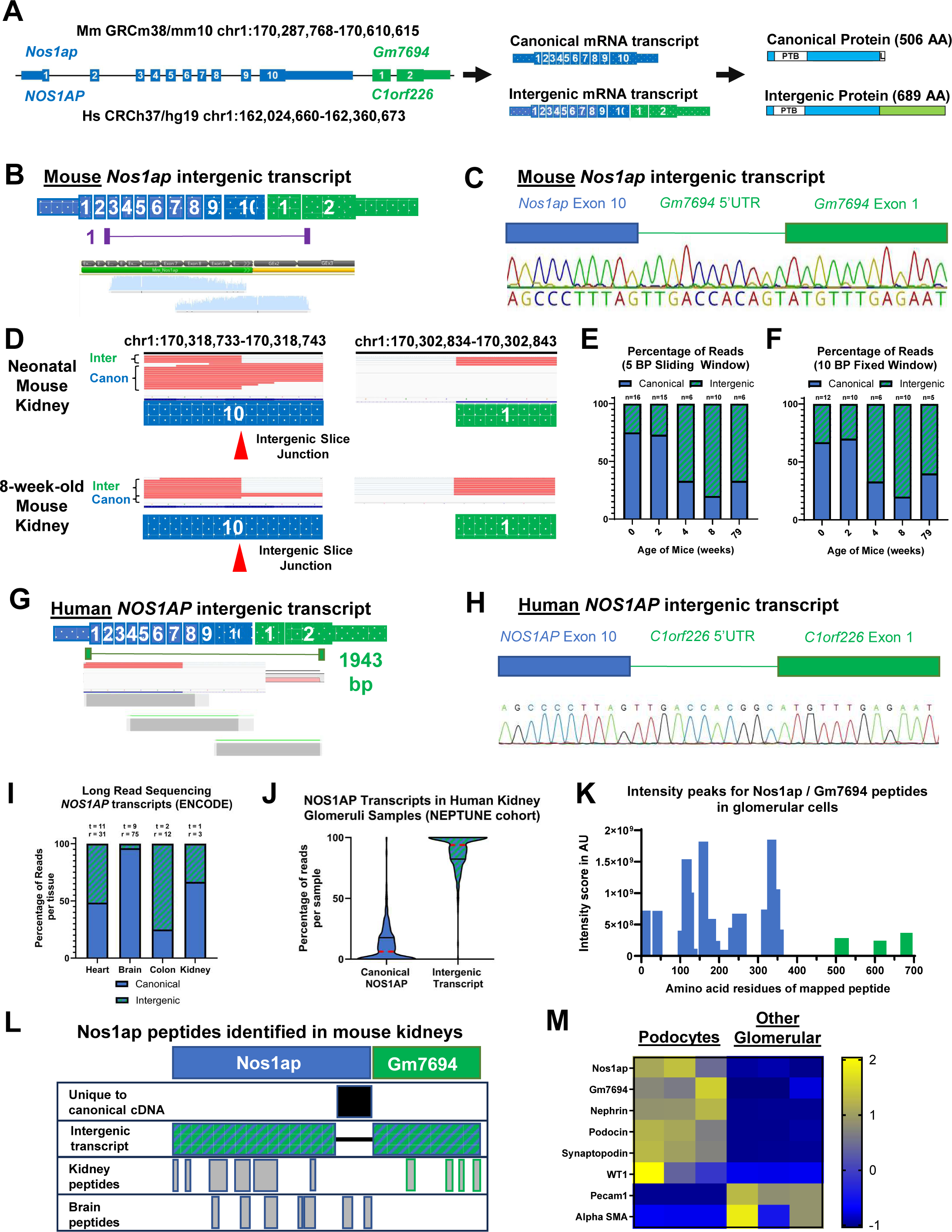
Detection of alternative *NOS1AP/Nos1ap* transcripts in mice and humans. (A) Cartoon depicting the NOS1AP/Nos1ap genomic locus in humans/mice is shown (left). Exons are indicated that contribute to the canonical and a non-canonical (intergenic) transcript. The latter results from intergenic splicing that joins 164 nucleotides from exon 10 of NOS1AP/Nos1ap to exons 1 and 2 of c1orf226/Gm7694. mRNA transcription and processing yield two potential transcripts (canonical and intergenic) seen in the middle diagram. Translation produces two potential protein products (right) containing the phospho-tyrosine binding (PTB) domain and, in the canonical protein, NOS1 PDZ binding domain (L). Exons/protein regions coded in blue contribute to both canonical and intergenic transcripts/isoforms while exons/protein regions coded in green only contribute to the intergenic isoform. (B) Cartoon depicting the murine *Nos1ap* intergenic splice product is shown. Exons are indicated that arise from the canonical *Nos1ap* (**blue**) or adjacent *Gm7694* (**green**) loci. The non-canonical transcript results from intergenic splicing that joins the first 153 nucleotides from exon 10 of *Nos1ap* to exons 1 and 2 of *Gm7694*. An amplicon spanning from early *Nos1ap* into exon 2 of *Gm7694*, which was identified by RT-PCR in adult mouse kidney total RNA. Aligned Sanger sequencing reads are shown below the transcript. (C) Sanger sequence reads of the PCR amplicon in (A) was aligned to the intergenic splice transcript, demonstrating a contiguous transcript from early exons in *Nos1ap* into *Gm7694* exon 2, including an in-frame 5’ UTR region of *Gm7694*. (D) 50 bp reads (red) from short read RNA sequencing of mouse kidneys were aligned to the *Nos1ap* and adjacent *Gm7694* loci. A greater fraction of canonical reads are shown extending through the intergenic splice junction in neonatal mouse kidney than in adult 8 week-old mouse kidney. (E) The ratio of canonical and intergenic reads in RNA sequencing data from 0-, 2-, 4-, 8-, and 79-week-old mice are shown using a 5 bp sliding window approach. Reads (n) are shown above box plot. (F) The ratio of canonical and intergenic reads is shown as in (E) using a 10 bp fixed window approach. Reads (n) are shown above box plot. (G) Cartoon is shown depicting the human *NOS1AP* intergenic splice product. Exons are indicated that arise from the canonical *NOS1AP* (**blue**) or adjacent *C1orf226* (**green**) loci. The non-canonical transcript results from intergenic splicing that joins the first 153 nucleotides from exon 10 of *NOS1AP* to exons 1 and 2 of *C1orf226*. The lower image shows aligned Sanger sequencing reads spanning nearly the full-size amplicon (1943 bp), which was identified by RT-PCR from adult human kidney tissue. This product starts within the 5’ UTR of *NOS1AP* and ends close to the stop codon within exon 2 of *C1orf226*. (H) Sanger sequencing plot resulting from the 1943 bp amplicon in (G) is shown demonstrating mRNA splicing between exon 10 of the canonical *NOS1AP* with the neighboring open reading frame *C1orf226*, including an in-frame 5’ UTR region of *C1orf226*. (I) Box plot displays percentage of *NOS1AP* transcripts reflecting canonical versus intergenic splice products by tissue in ENCODE long read RNA sequencing data. Tissues (t) and reads (r) are noted above the box plot. (J) Violin plot displays percentage of *NOS1AP* transcripts reflecting canonical versus intergenic splice products for each glomeruli sample in NEPTUNE cohort short read RNA sequencing data. Samples had minimum of 5 reads at the intergenic splice junction in *NOS1AP* exon 10. (K) Deep proteomics data from FACS-sorted podocytes and non-podocyte cells of mouse glomeruli was re-analysed with an amended list for sequences of non-canonical intergenic Nos1ap and Gm7694. Graph shows intensity peaks of all detected peptides mapped to respective amino acids residues of either Nos1ap or Gm7694. (L) Schematic overview showing rectangles representing either Nos1ap or Gm7694 or protein sequences (row 1), sequence unique two canonical Nos1ap (row 2, black rectangle), sequence of non-canonical intergenic product (row 3), mapping of the identified peptides in podocytes to their respective positions (row 4), mapping of the identified peptides in mouse brain tissue to their respective positions (row 5). (M) Heatmap comparing mean intensity scores for Nos1ap and Gm7694 as well as multiple glomerular marker proteins. Heatmap based on z-scores. Nephrin, podocin, synaptopodin and WT1, podocytic markers; Pecam1, endothelial cell marker; Alpha smooth muscle antigen, mesangial cell marker.

By RT-PCR of total RNA from 8-week-old mouse C57BL/6 kidneys, both contiguous canonical (blue only) and intergenic *Nos1ap* transcripts (blue and green) were detected (**Figure 1B-C, S2A-D**). The intergenic splice product arises from joining 153 nucleotides from exon 10 of *Nos1ap* to exons 1-2 of predicted gene *Gm7694* (**Figure 1A, 1C, S1**). To quantify the relative abundance of these isoforms, re-analysis of bulk short-read RNA sequencing data was performed from neonatal to aged C57BL/6 mouse kidneys (8-10 mice per group spanning 0 to 79 weeks of life). The ratio of intergenic to canonical reads was determined by their alignment to the canonical or intergenic transcript within an inclusive 5 bp sliding window or stringent 10 bp fixed window around the splice junction (GRCm38/mm10 chr1:170,318,737-170,318,738) (**Figure 1D, S3A-B**). Analyzed reads ranged from 5-16 per age group at this position out of 258.5-365.9 million aligned reads per age group. Canonical reads were more frequent at 0 and 2 weeks of life (67-75%; **Figure 1D-F, S3C**). In contrast, the intergenic reads were more frequent at 4, 8, and 79 weeks of life (60-80%, **Figure 1D-F, S3C**), suggesting an age-dependent regulation of this splicing event. The higher prevalence of intergenic reads was, furthermore, confirmed in an independent RNA sequencing dataset from 8-week-old C57BL/6 mice (**Figure S3D**).

*NOS1AP* intergenic splicing was, similarly, observed at the identical orthogonal splice junction (GRCh37 chr1:162336994-162336995) in adult human kidney tissue by RT-PCR (**Figure 1G-H, S2E**). Of note, this transcript did not include optional 15 nucleotides adjacent to exon 4 that have been detected in canonical human *NOS1AP* transcripts (**Figure S2E**). To examine contiguous *NOS1AP* transcripts in adult kidney or other tissues (age of acquisition 40 to >90 years), long read sequencing from the ENCODE project was interrogated and demonstrated contiguous intergenic transcripts in adult heart (n=11), brain (n=9), colon (n=2) and kidney (n=1) specimens (**Figure 1I, S4A-B**) to varying degrees relative to contiguous canonical transcripts. For example, the canonical *NOS1AP* transcript predominated across brain tissues (96%) while the ratio of canonical to intergenic transcript was more variable in heart tissues (48.4%) (**Figure 1I, S4A-B**).

As low read count in the single kidney sample from ENCODE only allowed for a qualitative analysis, we next performed a quantitative evaluation of short-read RNA sequencing data from human kidney samples in the NEPTUNE cohort (living donor biopsy, nephrectomy, and NS patient biopsy) covering 244 samples of individuals from 2 to >90 years. Quantifying reads that crossed the human splice junction in a 10 bp fixed window, we found that qualifying reads corresponding to the intergenic splice product predominated relative to canonical reads (69.6-93.8%) in all sample groups within RNA isolated from either glomerular or tubular compartments (**Figure 1J, S4C-D**).

Finally, to assess if peptides corresponding to either canonical or intergenic isoforms are abundant in glomerular podocytes, we re-analyzed proteomics data from FACS-sorted primary mouse glomerular cells (*29*). 23 unique peptides specific to either Nos1ap or Gm7694 were identified, predominantly in podocytes (**Figure 1K-M, Supp. File S1)**. Interestingly, no peptides aligning to the unique C-terminus of canonical Nos1ap were identified, despite theoretical trypsin cleavage sites yielding at least five detectable peptides of 7 to 25 amino acids in length (**Document S2**). No peptides spanning the junction from Nos1ap to Gm7694 could be detected either (**Document S2**). However, the only predicted digestion product including the Nos1ap-Gm7694 junction site has a peptide length of 54 amino acids, precluding detection by mass spectrometry (**Document S2**). In contrast, in proteomics data from adult mouse brain peptides mapping to the unique C-terminus of canonical Nos1ap but not Gm7694 were detected (**Figure 1L, Supp. File S2**) (*30*). This analysis provides evidence for the existence of Gm7694 on a protein level in glomerular podocytes.

Taken together, these observations indicate that the intergenic *NOS1AP* product is present in human and mouse kidneys and may be regulated in an age-dependent fashion.

### Variants impacting the *Nos1ap* locus cause murine podocytopathy

We, previously, described the kidney phenotype of *Nos1ap^Ex3-/Ex3-^* mice on a C57BL/6 background (*20*, *31*). These mice bear a homozygous deletion of the *Nos1ap* exon 3, causing an in-frame deletion within the N-terminal PTB domain (**Figure 2A, S1**). *Nos1ap^Ex3-/Ex3-^* mice developed moderate albuminuria (1-3 g albumin/g creatinine) accompanied by podocyte foot process effacement at an ultrastructural level. Because we previously reported a limited number of human subjects and mouse models (*20*), we characterized two novel *Nos1ap* mouse models to further establish the association between this locus and NS as well as to evaluate the biological relevance of the intergenic product.

**Figure 2.**
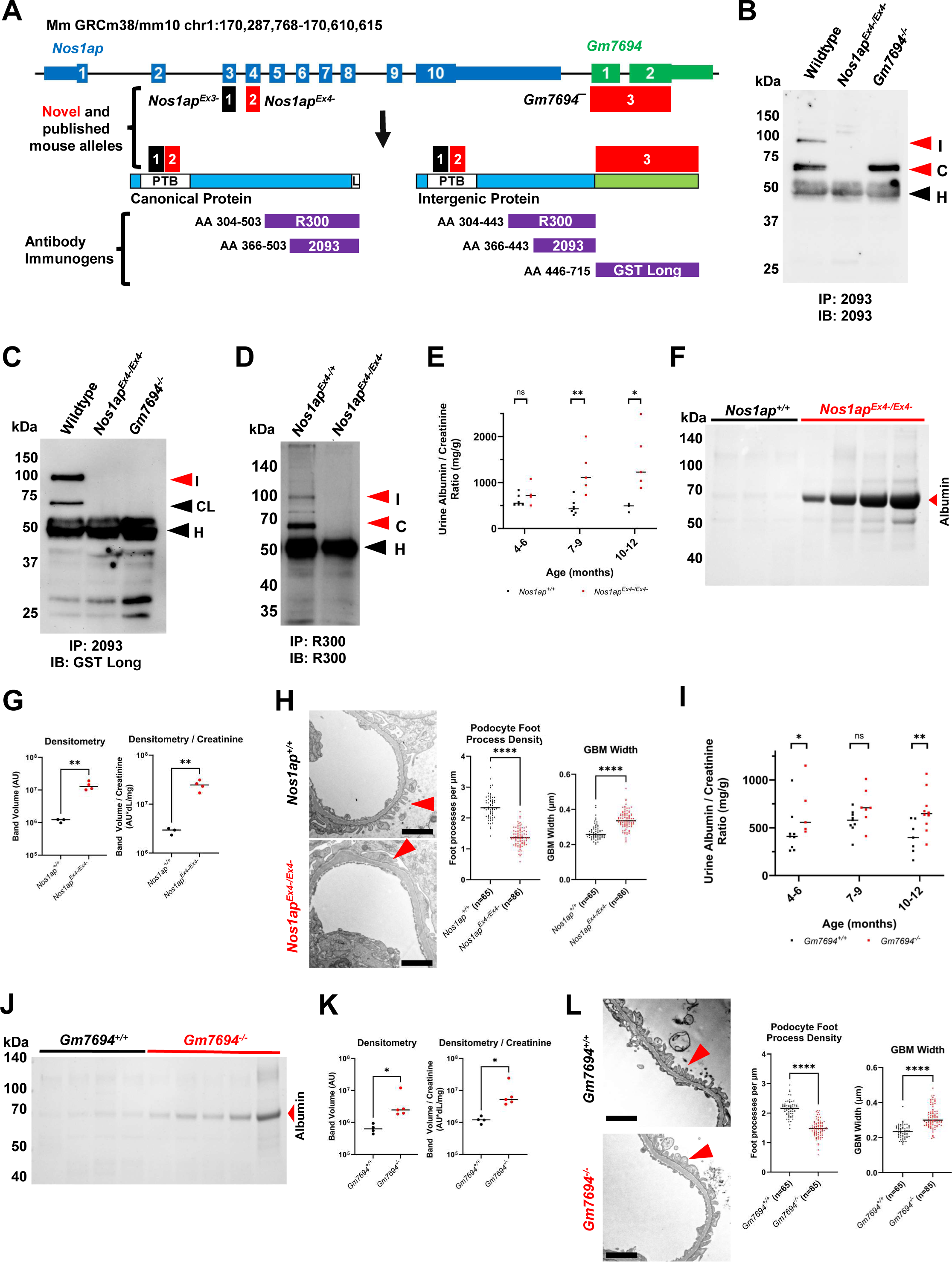
*Gm7694^-/-^* and *Nos1ap^Ex4-/Ex4-^* mice develop podocytopathies. (A) Cartoon is shown depicting the Nos1ap genomic locus in mice and murine alleles (black and red bars). Exons are indicated, which contribute to the canonical and non-canonical isoforms. The protein products are shown below with bars indicating the positions of regions deleted by the murine alleles. The two potential protein products (right) contain the phospho-tyrosine binding (PTB) domain, while only the canonical protein has a NOS1 PDZ binding domain (L). The positions of immunogens against which antibodies were raised are shown below (purple bars). (B) Immunoprecipitation (IP) using a rabbit polyclonal antibody (“2093”) raised against Nos1ap was performed in mouse embryonic fibroblasts isolated from wildtype, *Nos1ap^Ex4-/Ex4-^* and *Gm7694^-/-^* mice. Immunoblotting was performed on IP eluates using the same antibody and showed multiple protein bands consistent with Nos1ap isoforms in the wildtype eluate. Both Nos1ap isoform bands were absent in *Nos1ap^Ex4-/Ex4-^* fibroblast eluates. A band consistent with the canonical isoform remained in *Gm7694^-/-^* fibroblast eluate. (I: Intergenic Nos1ap isoform; C: canonical Nos1ap isoform; H: IgG Heavy Chain Band). (C) IP was performed as in (B). Immunoblotting was performed on IP eluates using an antibody specific to the intergenic Nos1ap isoform (“GSTlong”) and showed multiple protein bands consistent with the full-length intergenic isoform (I), a potential cleavage product of the intergenic isoform (CL) and the IgG Heavy Chain band (H) in the wildtype eluate. Nos1ap intergenic isoform bands were absent in both mutant fibroblast eluates. (D) Immunoprecipitation (IP) using a rabbit polyclonal antibody raised against Nos1ap (“R300”) was performed in neonatal mouse kidney lysates from wildtype and *Nos1ap^Ex4-/Ex4-^* mice. Nos1ap immunoblotting was performed on IP eluates and showed multiple protein bands consistent with Nos1ap isoforms that were absent in mutant kidney IP eluates. (I: Intergenic Nos1ap isoform; C: canonical Nos1ap isoform; H: IgG Heavy Chain Band). (E) Urine samples serially collected from 4-12-month-old *Nos1ap^Ex4-/Ex4-^* mice (n=5) show ACR levels that are significantly elevated relative to control mice (n=7). Graph shows dot plots with median bars for each genotype and time-point. Mann-Whitney test performed to compare groups at each timepoint (**p<0.01, *p<0.05, ^ns^p≥0.05 (F) Urine samples from 10-12-month old *Nos1ap^Ex4-/Ex4-^* mice and wildtype controls were interrogated by SDS-PAGE and Coomassie staining. *Nos1ap^Ex4-/Ex4-^* mice exhibited marked albuminuria (69 kDa protein band) above physiologic levels observed in wildtype mice. (G) Densitometry of albumin bands in (C) was performed and quantified, showing significantly increased abundance in 10-12-month old *Nos1ap^Ex4-/Ex4-^* mouse urine samples (n=4) relative to control mouse urine (n=3). Albumin protein band abundance normalized to urine creatinine was also significantly increased in *Nos1ap^Ex4-/Ex4-^* mouse urine samples relative to control mouse urine. Student’s t-test; **, p<0.01. (H) Representative transmission electron microscopy images are shown for *Nos1ap^Ex4-/Ex4-^* mice (n=4) and control wildtype mice (n=3). Red arrowheads point to tertiary foot processes. Semi-quantification of podocyte foot process density per µm of glomerular basal membrane (GBM) and quantification of GMB thickness in µm (left and right panel respectively). Scale bar 2 μm. (I) Urine samples serially collected from 4-12-month old *Gm7694^-/-^* mice (n = 10) show ACR levels that not significantly different to those of control mice urine samples (n = 12). Graph shows dot plots with median bars for each genotype and time-point. Mann-Whitney test performed to compare groups at each timepoint (**p<0.01, *p<0.05). (J) Urine samples from 10-12-month old *Gm7694^-/-^* mice (n=5) and wildtype controls (n=4) were interrogated by SDS-PAGE and Coomassie staining. Homozygotes exhibited albuminuria (69 kDa protein band) above physiologic levels observed in wildtype mice. (K) Densitometry of albumin bands in (B) was performed and quantified, showing significantly increased abundance in 10-12-month old *Gm7694^-/-^* mouse urine samples (n=5) relative to control mouse urine (n=4). Albumin protein band abundance normalized to urine creatinine was also significantly increased in *Gm7694^-/-^* mouse urine samples relative to control mouse urine. (L) Representative transmission electron microscopy images are shown for *Gm7694^-/-^* mice (n = 4) and control wildtype mice (n = 3). Red arrowheads point to tertiary foot processes. Semi-quantification of podocyte foot process density per µm of glomerular basal membrane (GBM) and quantification of GMB thickness in µm (left and right panel respectively). Scale bar 2 μm.

We first studied an additional mouse model, affecting both canonical and intergenic transcripts. *Nos1ap^Ex4-/Ex4-^* mice bear a homozygous out-of-frame deletion in *Nos1ap* exon 4, predicted to cause early truncation within the PTB domain (c.271_329del; p.91_110delfs*7) (**Figure 2A, S1**). Immunoprecipitation of endogenous Nos1ap from wildtype and *Nos1ap^Ex4-/Ex4-^* embryonic fibroblasts and neonatal mouse kidneys was performed using a polyclonal antibody raised against the Nos1ap C-terminus (**Figure 2A-B, 2D**). Immunoblotting for Nos1ap in the wildtype eluate revealed multiple protein bands between 50 and 100 kDa, which were absent in the *Nos1ap^Ex4- /Ex4-^* eluate (**Figures 2A, 2C**). Consistent with orthologous isoforms identified in primary rat neurons (*23*, *26*) and with mammalian kidney isoforms (**Figure 1**), the two most abundant bands exhibited sizes consistent with the canonical Nos1ap protein (∼70 kDa) as well as an intergenic Nos1ap-Gm7694 isoform (∼100 kDa) (**Figures 1A-C, 2A-B, 2D, S1**). Overall, this suggested the *Nos1ap^Ex4-^* allele causes total loss of Nos1ap protein expression in kidney tissue as expected.

Albuminuria was assessed in *Nos1ap^Ex4-/Ex4-^* mice between 4-12 months of age. Urine albumin-to-creatinine ratios (ACR) were significantly elevated in homozygous mice relative to wildtype controls at ages 7-9 months and 10-12 months (median ACR 1227 mg/g creatinine versus 492 mg/g in controls at 10-12 months) (**Figure 2E**). Analysis of urine samples from 10 to 12-month-old mice by SDS-PAGE and Coomassie staining revealed albuminuria across all homozygotes that was significantly elevated above the basal physiologic levels observed in control mice using band densitometry and normalization to urine creatinine (**Figure 2F-G**). Kidneys from approximately 10 to 12-month-old mice were evaluated for ultrastructural changes at the glomerular filtration barrier by electron microscopy. *Nos1ap^Ex4-/Ex4-^* mouse kidney sections exhibited reduced podocyte foot process density (median 1.36 foot processes/µm versus 2.34 in controls) and glomerular basement membrane thickening (median 335 nm versus 257 nm in controls) (**Figure 2H**). Overall, these observations supported that this recessive variant is associated with a murine podocytopathy, consistent with our published observations in *Nos1ap^Ex3- /Ex3-^* mice (*20*, *31*).

Next, we investigated an allele that only affects the intergenic splice isoform but not the canonical *Nos1ap* transcript. Specifically, we generated *Gm7694^-/-^* mice, which bear a complete deletion of predicted gene *Gm7694* (**Figure 2A, S1**). Immunoprecipitation of endogenous Nos1ap from embryonic fibroblasts showed selective loss of the intergenic but not canonical isoform in *Gm7694^-/-^* fibroblast eluates when immunoblotting with an antibody raised against rat Nos1ap C-terminal regions of the canonical and intergenic proteins (**Figure 2A-B**). Loss of the ∼100 kDa intergenic isoform was confirmed by immunoblotting of these lysates with an antibody raised selectively against the rat intergenic Nos1ap isoform (**Figure 2A, 2C**). This immunoblot also revealed a potential cleavage product between 50-75 kDa that was also absent in *Gm7694^-/-^* eluates (**Figure 2C**).

Albuminuria was assessed, showing significantly increased ACR levels in *Gm7694^-/-^* mice relative to controls between 4-6 months and 10-12 months (**Figure 2I**) (median ACR 647 mg/g creatinine in *Gm7694^-/-^* mice versus 397 mg/g in controls at 10-12 months). Analysis of urine samples from 10 to 12-month-old mice by SDS-PAGE and Coomassie staining similarly demonstrated albuminuria in homozygotes, which showed modest but significantly elevated above basal levels observed in control mice by densitometry and normalization to urine creatinine (**Figure 2J-K**). Kidneys from approximately 10- to 12-month-old mice were evaluated for ultrastructural changes by electron microscopy. Similar to *Nos1ap^Ex4-/Ex4-^* mice, *Gm7694^-/-^* mouse kidney sections exhibited reduced podocyte foot process density (median 1.47 foot processes/µm versus 2.16 in controls) and glomerular basement membrane thickening (median 300 nm versus 234 nm in controls) (**Figure 2L**). These findings suggest that *Gm7694* and the intergenic *Nos1ap* splice product play an important role in murine podocyte homeostasis and disease.

### Novel C-terminal recessive variants in *NOS1AP* are associated with human SRNS

We, previously, discovered that two recessive variants in *NOS1AP* cause early onset NS in humans (*20*). These variants were predicted to impact the N-terminal PTB domain of NOS1AP (**Figure S1, Table 1**). To discover novel disease-causing recessive variants in *NOS1AP* or the adjacent *C1orf226* loci in humans, exome data generated after the initial study was interrogated from an additional 985 families with NS. This revealed a rare, homozygous splice variant (NM_014697:c.1105+5G>C) in *NOS1AP* in 1 family (**Table 1**; **Figure 3, S1**). No competing variants were detected in additional 60 nephrotic syndrome disease genes or 11 monogenic nephritis disease genes (*7*). The subject is male and of Middle East descent, born from a consanguineous union. He developed edema during 0-5 years of age. Consistent with the clinical diagnosis of nephrotic syndrome, his initial laboratory evaluation revealed microscopic hematuria, nephrotic-range proteinuria (19 g protein/day), hypoalbuminemia (2.2 g/dL), reduced serum total protein levels (4 g/dL), and elevated serum triglyceride levels (9.98 mmol/L). His proteinuria was resistant to corticosteroids but partially responsive to the calcineurin inhibitor cyclosporine. Sanger sequencing confirmed the variant was present homozygously in the proband and heterozygously in unaffected parents and siblings, consistent with a recessive mode of inheritance (**Figure 3**). The variant was predicted to strongly impair splicing of the penultimate exon 9 by four independent algorithms (**Table 1; Figure S1**). Skipping of exon 9 is predicted to create an out-of-frame deletion and loss of NOS1AP or NOS1AP-C1orf226 C-terminal domains (**Figure S1**). Furthermore, the variant was absent from control genome databases ExAC and gnomAD (**Table 1**). We, thus, concluded the recessive *NOS1AP* variant identified in this family is deleterious and the likely cause of SRNS in this subject, highlighting the importance of the NOS1AP C-terminus in human podocyte homeostasis.

**Table 1.** Recessive mutations in *NOS1AP* (NM_014697) in families with nephrotic syndrome.

| Family<br>_Individual | Nucleotide<br>Change | Amino<br>Acid<br>Change | Exon<br>(Zyg,<br>Seg) | In silico<br>Severity<br>Scores | Conser-<br>vation | ExAC<br>and<br>gnomAD | S<br>E<br>X | Ethnic<br>origin | PC | Renal Disease |
| --- | --- | --- | --- | --- | --- | --- | --- | --- | --- | --- |
| Family 1 | c.1105+5G>C | Splice | 9<br>(hom, Y) | SSF -100%<br>ME -100%<br>NNS -100%<br>GS -100% | N/A | NP | M | Syria | Y | <u>Initial Onset</u>: 3 years, edema<br><u>Urinalysis</u>: Microscopic hematuria; 19 g protein/day<br><u>Serum studies</u>: Cr 0.23 mg/dL, TP 4 g/dL, Albumin 2.2 g/dL, TG 9.98 mmol/L<br><u>Biopsy</u>: FSGS<br><u>Treatment</u>: SRNS, Partial response to CsA. |
| Family 2<br>(Clinvar) | Canonical<br>c.1259G>C<br>Intergenic<br>c.1258+1G>C | p.G420A<br>Splice | 10<br>(hom) | SIFT t<br>MT b<br>PP2 b<br>SSF -100%<br>ME -100%<br>NNS -100%<br>GS -100% | X. tropicalis<br>N/A | NP | M | Saudi<br>Arabia | Y | <u>Initial Onset</u>: Congenital NS<br><u>Renal function</u>: ESKD at age 3 years<br><u>Course</u>: Passed away at age 5 years<br><u>RUS</u>: Unilateral multicystic kidney |
| Family3*<br>(published) | c.428G>A | p.C143Y | 5 (hom,<br>mat) | SIFT DL<br>MT DC<br>PP2 PD | C. elegans | NP | M | NA | Y | <u>Initial Onset</u>: 4 days, SGA, edema<br><u>Urinalysis</u>: Microscopic hematuria; 18.5 g protein/g creatinine<br><u>Serum Studies</u>: Cr 1.0 mg/dL (CKD), TP 4.8 g/dL.<br><u>RUS</u>: N/A<br><u>Treatment</u>: Resistant to corticosteroids, cyclophosphamide, cyclosporin A.<br><u>Biopsy</u>: Mesangial cell proliferation, 75% podocyte foot effacement.<br><u>ESKD</u>: 7 years (HD and TX). |
| Family4*<br>(published) | c.345-3T>G | Splice | 5 (hom) | SSF -5%<br>ME -66%<br>NNS -100%<br>GS -77% | N/A | NP | M | Egypt | Y | <u>Initial Onset</u>: 6 months, edema<br><u>Urinalysis</u>: Microscopic hematuria; 3 g protein/g creatinine.<br><u>Initial Renal Function</u>: Cr 0.6 mg/dL (CKD).<br><u>RUS</u>: Bilateral echogenicity<br><u>Treatment</u>: Resistant to corticosteroids. |

Abbreviations: CsA, cyclosporine; CKD, chronic kidney disease; Cr, serum creatinine; DC, disease causing; DL, deleterious; ESKD, End-stage Kidney Disease; ExAC v1.0 and gnomAD v2.11 and v3, summary of data within Exome and Genome Aggregation databases; FSGS, focal segmental glomerulosclerosis; HD, hemodialysis; hom, homozygous; ME, MaxEnt splice prediction score; NA, not available; N/A, not applicable; NS, nephrotic syndrome; NNS, NNSPLICE splice-site mutation prediction score; PC, parental consanguinity; PD, probably damaging; PP2, PolyPhen-2 prediction score; RUS, renal ultrasound; SGA, small for gestational age; SIFT, "Sorting Tolerant From Intolerant" prediction score; Seg, segregation; SRNS, steroid-resistant nephrotic syndrome; TG, triglycerides; TP, total protein; TX, kidney transplantation; Y, yes; Zyg, zygosity. \*Published in Majmundar, Buerger et al., *Sci Adv* 2021.

**Figure 3.**
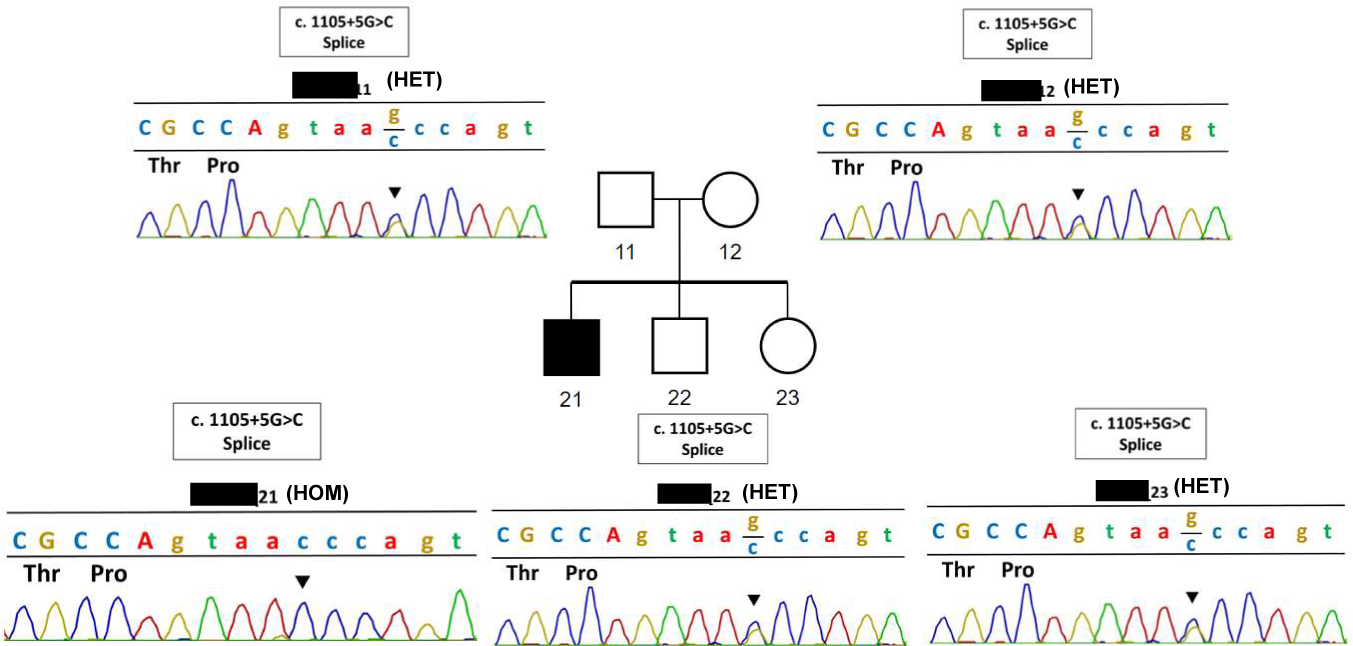
Novel recessive variant in *NOS1AP* associated with pediatric SRNS. Pedigree of family members shown. Shaded symbols indicate the individual has SRNS while open symbols indicate unaffected status. Sanger tracings for affected subject _21 carries the homozygous *NOS1AP* variant c.1105+5G>C while parents and unaffected siblings are heterozygous at this position.

In addition, we analyzed all *NOS1AP* or *C1orf226* variants in the ClinVar genomic variation database (*32*) for deleteriousness under the hypotheses that (i) recessive variants in these loci are associated with nephrotic syndrome and (ii) variants were misclassified based on the canonical transcript alone. Focusing on non-structural variants (<50 bp in size), we observed 67 reported variants, of which 14 variants were deemed deleterious based on rare prevalence and predicted impact on protein coding or splice effect. Of these 14 variants, we previously published two (**Figure S1, Table 1**) (*20*). We were unable to obtain further clinical genetic data for three novel variants. 8 additional variants were found in heterozygous states. A final variant was observed in a subject (ClinVar family 2), where detailed genotype and phenotype data was available. Exome sequencing had been performed, identifying a homozygous variant in *NOS1AP* exon 10 that was absent from the gnomAD database (**Table 1**). This single nucleotide variant was predicted to cause a missense in the canonical product (NM_014697: c.1259G>C; p.G420A) with weak *in silico* prediction scores and, therefore, was not deemed deleterious. However, in context of the alternative *NOS1AP-C1orf226* transcript, the variant alters the +1 position of the intergenic splice site within *NOS1AP* exon 10 (c.1258+1G>C). Multiple prediction scores strongly indicate impaired splicing and thus deleteriousness. Of note, no competing variants were detected, including in >71 genes associated with Mendelian genetic nephrotic syndrome and/or nephritis (*7*). Interestingly, reverse phenotyping of the subject revealed the most severe NS phenotype, congenital nephrotic syndrome, which had later progressed to end-stage kidney disease requiring dialysis. The subject had passed away within the age range of 0-5 years. He also had a unilateral cystic kidney and healthy appearing contralateral kidney, a condition unlikely to be responsible for his early onset kidney failure. This recessive *NOS1AP* variant highlights that intergenic splicing of *NOS1AP* and *C1orf226* is critical in human podocyte homeostasis and corroborated our findings in mouse model studies (**Figure 2**).

### Recessive Nos1ap variants cause severe albuminuria and kidney dysfunction on FVB/N genetic background

While we previously demonstrated that *Nos1ap^Ex3-/Ex3-^* mice on a C57BL/6 background developed proteinuric disease, these mice did not exhibit persistent hypoalbuminemia, kidney dysfunction nor reduced survival (*20*)—features frequently observed in human SRNS. Moreover, the novel mouse models described here similarly had modest albuminuria and normal survival into late adulthood on C57Bl/6 background (**Figure 2**). Because genetic background can modify the severity of murine podocytopathies (*33–35*), we posited that the kidney phenotype in C57BL/6- *Nos1ap^Ex3-/Ex3-^* could be similarly affected by genetic background to faithfully recapitulate human SRNS.

C57BL/6-*Nos1ap^Ex3-/+^* mice were, thus, bred with FVB/N or 129/sv mice for more than six generations. This would yield *Nos1ap^Ex3-/+^* mice with a genetic background reflecting >98% of the outcrossed strain. Resulting FVB/N-*Nos1ap^Ex3-/+^* or 129/sv-*Nos1ap^Ex3-/+^* were incrossed to yield wildtype, heterozygote, and homozygote mice of the same background. FVB/N-*Nos1ap^Ex3-/Ex3-^* mice developed albuminuria at weaning age with median ACR of 1.9 g/g, that increased to 13.8-15.7 g/g at 4-6 months of life (**Figure 4A**). In contrast, ACR levels in 129/sv-*Nos1ap^Ex3-/Ex3-^* mice were more modestly elevated relative to wildtype or heterozygote littermate controls (1-3 g/g during the first six months of life) (**Figure S5A**). Overall, FVB/N-*Nos1ap^Ex3-/Ex3-^* mice exhibited markedly elevated albuminuria relative to 129/sv-*Nos1ap^Ex3-/Ex3-^* and C57BL/6-*Nos1ap^Ex3-/Ex3-^* mice between 3-6 months of life (**Figure 4B**), indicating the FVB/N background uniquely modifies this kidney phenotype.

**Figure 4.**
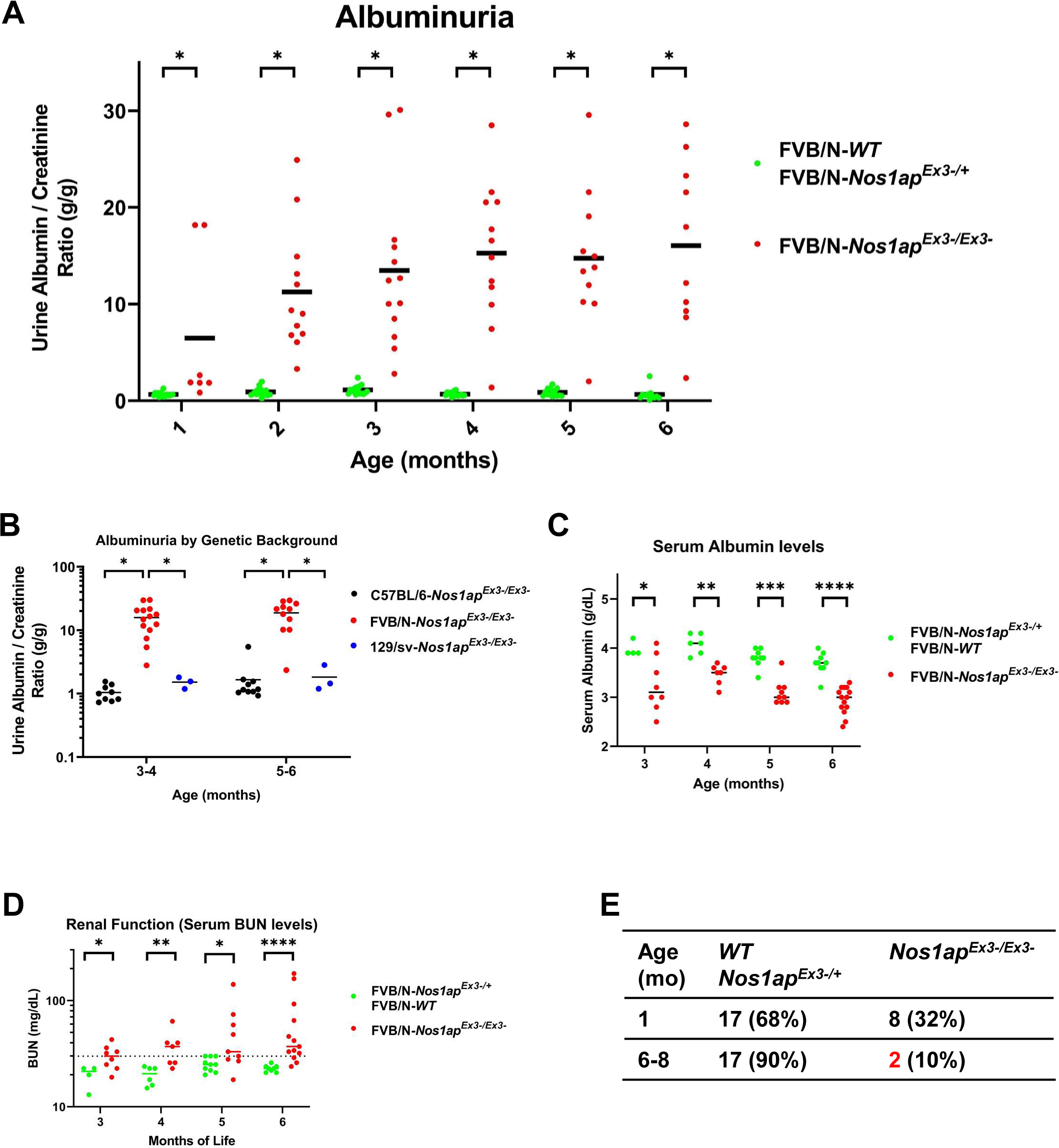
FVB/N-*Nos1ap^Ex3-/Ex3-^* mice develop early-onset progressive albuminuria. (A) *Nos1ap^Ex3-/Ex3-^*mice on an FVB/N background show significantly elevated urine albumin-to- creatinine ratios (ACR) beginning at weanling age and increasing through 4-5 months of age compared to control animals. *Nos1ap^Ex3-/Ex3-^* group consists of 9 male and 7 female mice. Control mice consisted of 11 *Nos1ap^Ex3-/+^* heterozygous mice (6 male, 5 female) as well as 3 wildtype littermates (1 male, 2 female). Graph shows dot plots with mean bars for each genotype and time- point. Red, homozygous animals; Green, wild type and heterozygous littermate controls. Mann- Whitney test performed to compare groups at each timepoint (*p<0.05). (B) *Nos1ap^Ex3-/Ex3-^* on an FVB/N background (n = 14 mice) show ACR levels that are one order of magnitude above ACR levels detected in the originally studied C57BL/6 (n = 10 mice) as well as 129/sv background (n = 3 mice). Graph shows dot plots with mean bars for each genotype and time-point. Red, FVB/N-*Nos1ap^Ex3-/Ex3-^* ; Black, C57BL/6-*Nos1ap^Ex3-/Ex3-^* ; Blue, 129/sv-*Nos1ap^Ex3- /Ex3-^*. Mann-Whitney test performed to compare groups at each timepoint (*p<0.05). (C) Serum albumin levels were quantified using the VetScan VS2 system and “Comprehensive Diagnostic Profile” rotors. Homozygous animals (n = 17 mice) showed significantly reduced levels of serum albumin relative to control mice (n = 9 mice). Red, homozygous animals; Green, wild type and heterozygous littermate controls; mo, months. Mann-Whitney test performed to compare groups at each timepoint (****p<0.0001, ***p<0.001, **p<0.01, *p<0.05). (D) Serum BUN levels were quantified in homozygotes (red) or littermate controls (green) from 3- 6 months of life. A subset of homozygotes (17/29 measurements) showed elevated BUN levels above 30 mg/dL at 3-6 months of life while no control samples did (0/21 measurements). (****p<0.0001, **p<0.01, *p<0.05). (E) Animal numbers were assessed of FVB/N-*Nos1ap^Ex3-/Ex3-^* mice and littermate controls (*WT*, *Nos1ap^Ex3-/+^*), which underwent minimal interventions, at 1 month of life at time of weaning and at 6-8 months of life. Homozygotes showed increased mortality between 6-8 months of life that was not observed in littermate controls.

FVB/N-*Nos1ap^Ex3-/Ex3-^* mice were further analyzed for serum markers of nephrotic syndrome. In association with marked albuminuria, FVB/N homozygous mice developed significantly reduced serum albumin levels in serial measurements at 3-6 months of life relative to littermate controls (median albumin levels 3.0-3.5 g/dL versus 3.7-4.1 g/dL) (**Figure 4C**). Serum BUN levels, a marker of declining kidney function, at 3-6 months were significantly elevated in FVB/N- *Nos1ap^Ex3-/Ex3-^* mice relative to littermate controls (median BUN levels 30-37 mg/dL versus 20-25 mg/dL) (**Figure 4D, S5B**). In correlation with reduced kidney function, homozygotes also exhibited reduced survival between 6-8 months of life relative to control mice (**Figure 4E**). Thus, in contrast to C57BL/6-*Nos1ap^Ex3-/Ex3-^* mice (*20*), FVB/N-*Nos1ap^Ex3-/Ex3-^* mice develop persistent hypoalbuminemia, kidney dysfunction, and increased mortality, faithfully recapitulating features of human SRNS.

### FVB/N-*Nos1ap^Ex3-/Ex3-^* mice exhibit histologic and ultrastructural features of a severe podocytopathy

Given the urinary and serum abnormalities in FVB/N-*Nos1ap^Ex3-/Ex3-^* mice, we hypothesized homozygous mice develop glomerular changes consistent with a podocytopathy. Kidney tissue sections from 6-8 months old mice were Periodic Acid Schiff (PAS) stained and analyzed by light microscopy. Measurement of PAS-positive matrix within glomerular tufts revealed significantly increased mesangial matrix deposition, indicative of chronic glomerular injury, in FVB/N- *Nos1ap^Ex3-/Ex3-^* mice relative to heterozygote controls (**Figure 5A**). Kidney tissue sections from FVB/N homozygotes, furthermore, showed tubular dilation, reminiscent of human congenital nephrotic syndrome (*36*), in contrast to heterozygote controls (**Figure 5B**).

**Figure 5.**
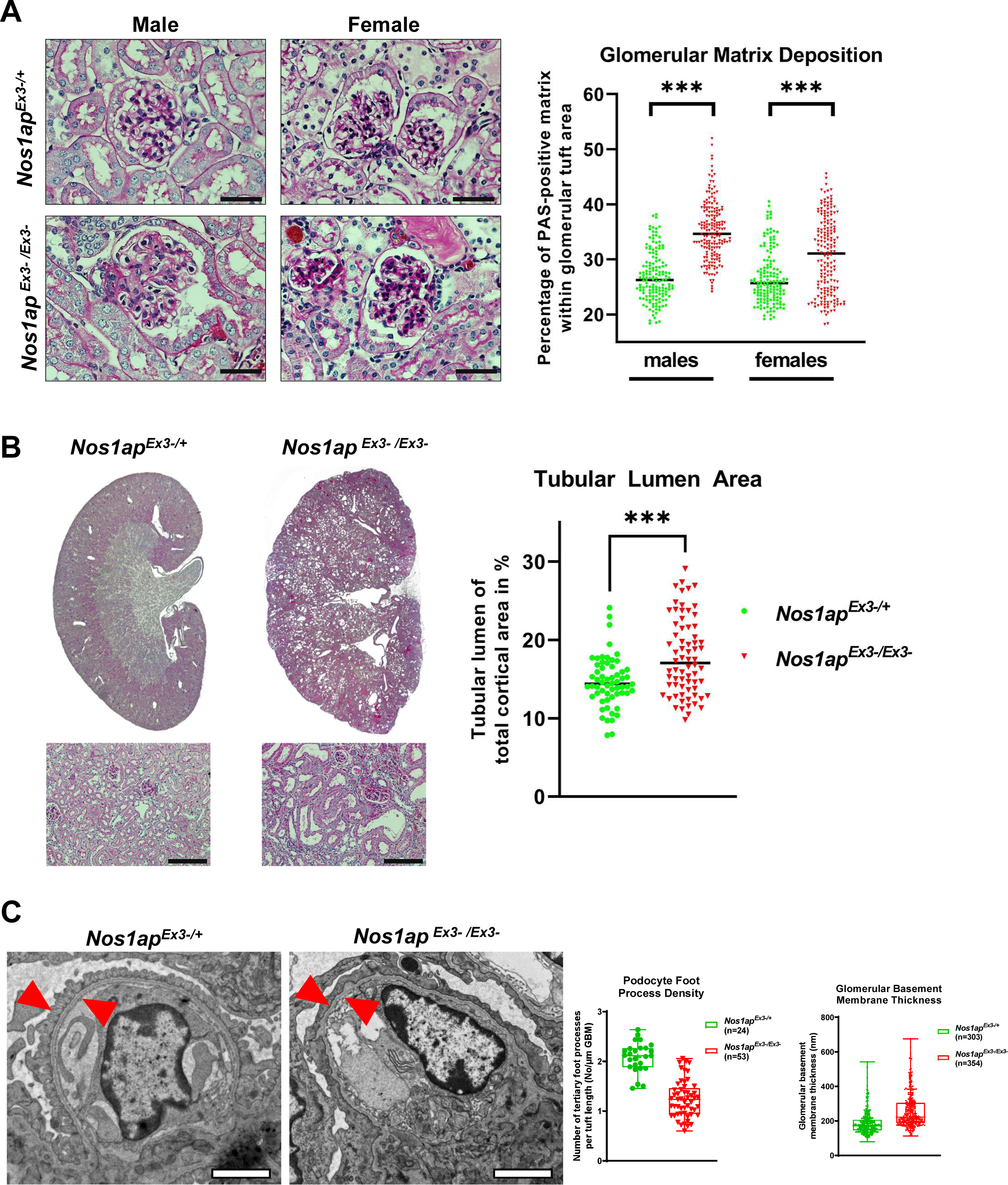
FVB/N-*Nos1ap^Ex3-/Ex3-^* mice develop glomerular matrix deposition, tubular dilation, and ultrastructural changes to the glomerular filtration barrier. (A) Periodic acid–Schiff stained sections of kidneys from 6 month old *Nos1ap^Ex3-/Ex3-^* and *Nos1ap^Ex3-/+^* mice were generated. 40x images of glomeruli were evaluated through an automated ImageJ pipeline to determine glomerular matrix deposition in a blinded manner. Both male and female *Nos1ap^Ex3-/Ex3-^* mice showed significantly increased glomerular matrix deposition. 6-7 mice (151-177 glomeruli) per genotype/sex group were analyzed. Mann-Whitney test; ***, p<0.001; Red, *Nos1ap^Ex3-/Ex3-^*; Green, *Nos1ap^Ex3-/+^* littermate controls; Scale bar μm. (B) Kidneys were processed at in (A). Representative overview images (stitched from 2X fields) and inset images (20X) are shown for *Nos1ap^Ex3-/+^* controls and *Nos1ap ^Ex3-/Ex3-^* mice. Tubular dilation is observed in homozygous mice as quantified in the dot plot (right). Red, *Nos1ap^Ex3-/Ex3-^*; Green, *Nos1ap^Ex3-/+^* littermate controls. n=12-14 mice per group, five 20X fields per mouse for quantification; Mann-Whitney test; ***, p<0.001. Scale bar 100 μm. (C) Representative transmission electron microscopy images are shown for the heterozygous control *Nos1ap^+/Ex3-^* and homozygous *Nos1ap ^Ex3-/Ex3-^* mice. Red arrowheads point to tertiary foot processes. Semi-quantification of podocyte foot process density per µm of glomerular basal membrane (GBM) and quantification of GMB thickness in nm (left and right panel respectively). Scale bar 2 μm.

Kidney sections were also examined by electron microscopy at weaning age (3-4 weeks of life) to assess early ultrastructural changes when albuminuria was first detected in FVB/N-*Nos1ap^Ex3- /Ex3-^* mice (**Figure 4A**). Homozygotes exhibited podocyte foot process effacement and thickening of the glomerular basement membrane (GBM) (**Figures 5C**), relative to heterozygote glomeruli exhibiting appropriate rhythmicity of foot process formation and GBM thickness. Overall, *Nos1ap^Ex3-/Ex3-^* mice exhibited biochemical and structural kidney abnormalities consistent with human SRNS and modified by genetic background.

### FVB/N-*Nos1ap^Ex3-/Ex3-^* mice respond to RAAS blockade

RAAS blockade is an established treatment in human Alport syndrome caused by genetic variants in the glomerular basement membrane related genes *COL4A3/4/5* (*37*). This treatment was initially validated in genetic mouse models of Alport syndrome (*38*). However, it remains unclear if this clinical benefit extends to other monogenic forms of nephrotic syndrome. This includes those caused by variants in podocyte-specific actin regulatory genes such as *NOS1AP* (*10*, *19*, *20*, *39–42*).

To evaluate this hypothesis, FVB/N-*Nos1ap^Ex3-/Ex3-^* mice were treated at weaning age with oral lisinopril. To assess the impact on proteinuria, urine ACRs were measured every two weeks for 14 weeks of treatment (**Figure 6A**). FVB/N-*Nos1ap^Ex3-/Ex3-^* mice treated with 100 mg/L and 200 mg/L lisinopril exhibited significantly reduced albuminuria relative to vehicle treated animals (**Figure 6A**), indicating RAAS ameliorates the proteinuric phenotype in this model.

**Figure 6.**
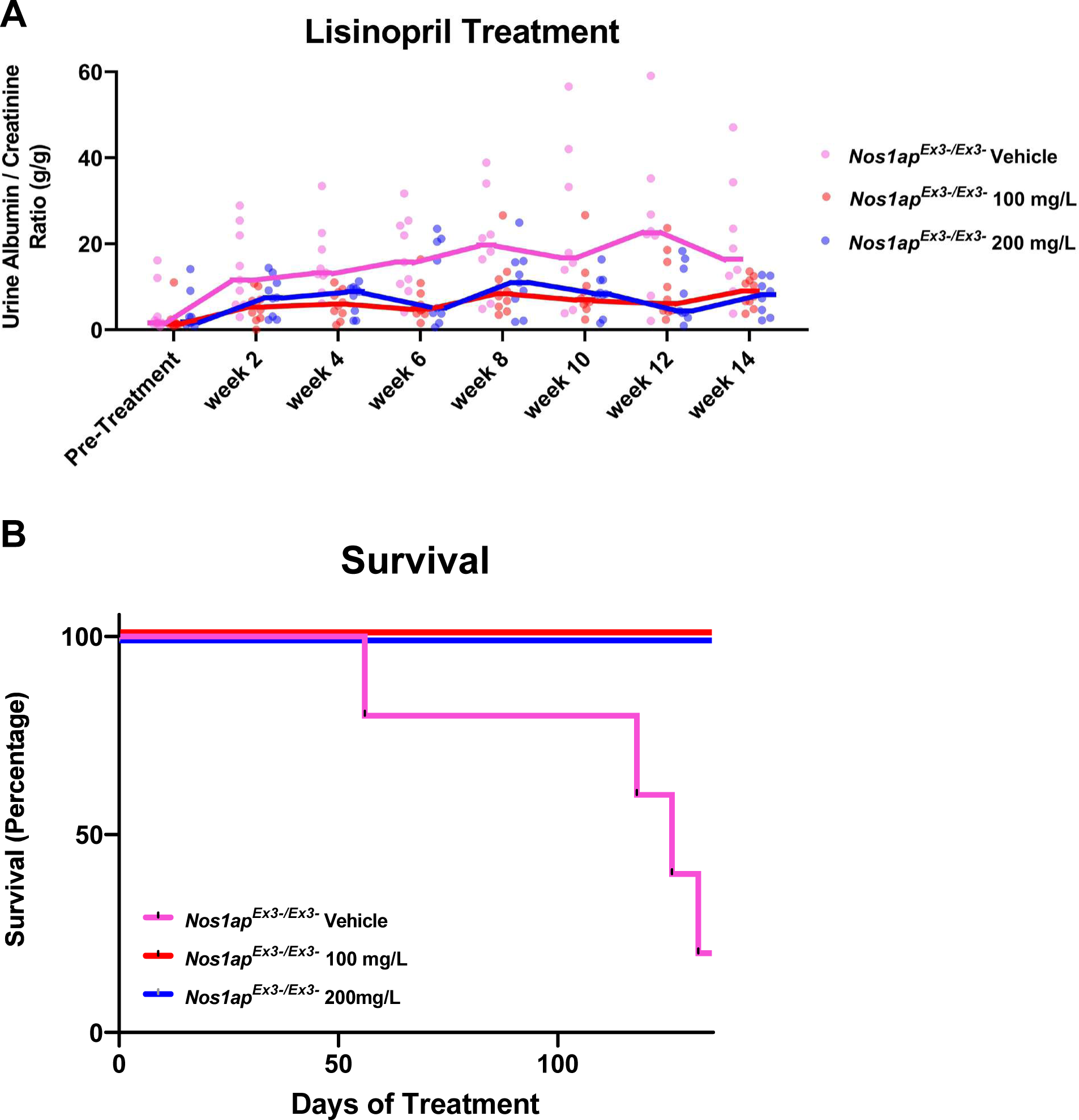
Anti-proteinuric treatment reduces albuminuria and prevents mortality in FVB/N-*Nos1ap^Ex3-/Ex3-^* mice. (A) FVB/N-*Nos1ap^Ex3-/Ex3-^*mice with comparable baseline ACRs received drinking water with either vehicle (water), 100 mg/L lisinopril, or 200 mg/L lisinopril starting at 6 weeks of age. Urine was collected biweekly and albumin as well as creatinine was quantified to determine albumin-to-creatinine ratios (ACR) as in Fig. 1. In both treatment groups (100 mg/L lisinopril or 200 mg/L lisinopril) albuminuria progression was significantly reduced by Kruskal-Wallis test (p<0.01 for vehicle versus 100 mg/L, p<0.05 for vehicle versus 200 mg/L). Pink, Vehicle control, n=9 (5 male, 4 female); Red, 100 mg/L lisinopril group, n=10 (5 male, 5 female); Blue, 200 mg/L lisinopril group, n=9 (4 male, 5 female). (B) FVB/N-*Nos1ap^Ex3-/Ex3-^*mice from (A) treated with lisinopril were followed up past urine collections for a total of 140 days of treatment. During the observed period none of the lisinopril-treated mice died (0/11) while only 20% of untreated mice survived (4/5 died). Pink, Vehicle control, n=5; Red, 100 mg/L lisinopril group, n=6; Blue, 200 mg/L lisinopril group, n=5. Statistically significant by Mantel-Cox test (p = 0.0014).

Serum markers were measured at early (4-8 weeks of treatment) and late (12-16 weeks of treatment) time-points. Serum albumin and total protein were significantly elevated in homozygous mice treated with 100 mg/L lisinopril relative to those receiving vehicle at the early timepoint (**Figure S6A-B**), correlating with the improvement in albuminuria (**Figure 6A**). FVB/N- *Nos1ap^Ex3-/Ex3-^* mice treated with 100 mg/L also showed reduced BUN levels relative to vehicle treated animals at the late timepoint (**Figure S6C**), indicating kidney dysfunction was blunted by this ACE inhibitor dose. In contrast, FVB/N-*Nos1ap^Ex3-/Ex3-^* mice treated with 200 mg/L did not have significantly different serum albumin, total protein, or BUN levels than vehicle treated-mice (**Figure S6**).

Survival of FVB/N-*Nos1ap^Ex3-/Ex3-^* mice was monitored over 140 days of treatment. While only 20% (1/5) of vehicle-treated mice survived this time frame, all (11/11) of the homozygotes treated with lisinopril survived (**Figure 6B**). These results indicate that RAAS blockade using lisinopril not only reduces proteinuria but also prevents mortality in a mouse model of human SRNS caused by defective actin remodeling.

### FVB/N-*Nos1ap^Ex3-/Ex3-^* mice do not respond to Dynamin Activator Bis-T-23

Dynamin stimulates actin bundling and, like NOS1AP, promotes filopodia formation (*20*, *23*, *43*–*45*). Dynamin activating compound Bis-T-23 reduced albuminuria and normalized podocyte ultrastructural defects in multiple murine podocytopathies (*46*). We hypothesized that Bis-T-23 would, similarly, improve proteinuria in FVB/N-*Nos1ap^Ex3-/Ex3-^* mice. However, we did not observe altered albuminuria with daily Bis-T-23 treatment in 4-week-old FVB/N-*Nos1ap^Ex3-/Ex3-^*mice including when collecting urine 1-3 hours after treatment to assess for transient reduction in albuminuria. (**Figure S7**).

## DISCUSSION

In summary, we demonstrate that novel variants impacting the *NOS1AP-C1orf226* locus can cause a podocytopathy (**Figure 1-3**), supporting an important role for the, here described, *NOS1AP* intergenic splice product in kidney disease. We, furthermore, demonstrate that genetic background can modify the severity of this genetic podocytopathy (**Figure 4-5**), which resulted in the generation of a more faithful model of human NS and suggested that variants in additional loci can modify the nephrotic syndrome trait in mammals. Lastly, we provide evidence that in a mouse model of *NOS1AP*-associated podocytopathy RAAS blockade is effective in reducing proteinuria and preventing early mortality in this genetic form of nephrotic syndrome (**Figure 6**).

The novel *NOS1AP* alleles in humans and mice identified in this study (**Figures 2-3**, **S1; Table 1**) not only expand the genotype-phenotype correlation between this locus and NS but also underscore the impact of C-terminal domains and alternative splice isoforms of *NOS1AP/Nos1ap* in monogenic nephrotic syndrome. It does remain unclear if the canonical and intergenic isoforms of NOS1AP play redundant or independent roles in podocyte biology.

The variant identified in ClinVar family2 was misclassified in the initial CLIA-certified result due to interpretation based on the canonical transcript. This case underscores the importance of tissue- and condition-specific isoform expression for correct variant interpretation, especially as recent studies indicate that >10 alternative transcripts exist for human coding genes while only an average of 4 transcripts per gene are annotated (*47–49*).

Our previous studies indicate that NOS1AP-dependent actin remodeling in podocytes is independent of its neuronal binding partner NOS1 (*20*). It remains possible that the canonical NOS1AP protein, which is abundant in neonatal mouse kidney, may mediate its actin remodeling effects through a key interaction partner other than NOS1. In addition, our current findings lead us to posit that NOS1AP may regulate podocyte homeostasis through interactions mediated by the novel C-terminal domain created through the intergenic splice product. Future studies should compare isoform-specific interaction partners and determine if in-frame deletion of the NOS1- binding domain causes a podocytopathy in mice.

Transcriptional and protein studies suggest predominance of the novel intergenic over the canonical transcript in human and mouse kidneys (**Figure 1, 2**). Limitations remain regarding (i) the relevance of the canonical isoform during kidney development and (ii) the prevalence of short *NOS1AP-C1orf226* transcripts not distinguished by short read RNA sequencing data.

Genetic background is, next, established to modify the phenotype of *Nos1ap^Ex3-/Ex3-^* mice and thereby yields a more faithful model of steroid-resistant NS (**Figures 3-4, S3**). In fact, the FVB/N- *Nos1ap^Ex3-/Ex3-^* mice develop profound albuminuria, kidney dysfunction, and histologic kidney damage reminiscent of human congenital nephrotic syndrome caused by genetic variants in *NPHS1* (*36*). The increased susceptibility of the FVB/N mouse strain to glomerular injury in genetic and acquired models of glomerular disease has been previously established (*34*, *35*, *50*–*53*). Genome-wide association studies have been performed but revealed distinctly associated loci depending on the kidney disease model (*35*, *50–52*). It will be important in future studies to identify loci specifically associated with modification of the *Nos1ap^Ex3-/Ex3-^* albuminuria and kidney dysfunction traits, as these genetic factors could reveal modifying factors that mitigate or aggravate the actin dysregulation caused by *Nos1ap* deficiency.

Treatments are limited in genetic forms of NS. RAAS blockade is widely employed in acquired proteinuric diseases providing reduction of proteinuria and preservation of GFR (*54–56*). Some case reports and a recent metanalysis report proteinuria reduction from RAAS blockade in children with genetic or primary SRNS/FSGS (*57–61*). However, strong evidence for clinical or experimental benefits in terms of preservation of renal function and reduction in mortality has only been demonstrated for *COL4A3/4/5*-variant associated Alport syndrome (*37*, *38*). Our findings that lisinopril ameliorates *Nos1ap-*associated podocytopathy in mice suggests that this benefit extends to other Mendelian genetic pathways and should be considered in other forms caused by variants in podocyte-specific actin regulatory genes (*10*, *19*, *20*, *39–42*).

On the other hand, Bis-T-23 did not alter the proteinuria in FVB/N-*Nos1ap^Ex3-/Ex3-^* mice despite the related roles that NOS1AP and dynamin proteins play in actin-based filopodia formation (*20*, *23*, *43–45*). It should be noted that there were, in fact, only modest transient effects of Bis-T-23 on proteinuria in a mouse model of *ACTN4*-associated nephropathy (*46*), indicating enhancing Dynamin oligomerization may not be sufficient to restore normal podocyte physiology in some forms of genetic NS.

Overall, these findings (i) underline the importance of understanding tissue-specific isoform expression for interpreting genetic testing results and (ii) further establish an essential role of NOS1AP for podocyte biology and podocytopathies.

## METHODS

### Study approval

Approval for human subjects research was obtained from Institutional Review Boards of the University of Michigan, Boston Children’s Hospital, and local IRB equivalents.

### Research subjects

The Hildebrandt laboratory obtained blood samples and pedigrees following informed consent from individuals with NS or their legal guardians. The diagnosis of NS was based on clinical features of nephrotic-range proteinuria, hypoalbuminemia, and edema and supported by kidney biopsy findings evaluated by renal pathologists. Clinical data were obtained using a standardized questionnaire (http://www.renalgenes.org).

The Al-Hamed laboratory recruited subjects with informed consent for genetic studies under IRB- approved research protocols RAC#2160022 at King Faisal Specialist Hospital and Research Centre (KFSH&RC).

### ES, and variant calling

Exome sequencing (ES) and variant calling were performed as previously described(*7*, *62*) to discover a novel genetic cause of NS using Agilent SureSelect™ human exome capture arrays (Thermo Fisher Scientific) with next generation sequencing (NGS) on an Illumina™ platform. Sequence reads were mapped against the human reference genome (NCBI build 37/hg19) using CLC Genomics Workbench (version 6.5.1) (CLC bio). Genetic location information is according to the February 2009 Human Genome Browser data, hg19 assembly (http://www.genome.ucsc.edu). Downstream processing of aligned BAM files were done using Picard and samtools, and SNV calling was done using GATK5. Variant calling was performed as previously described(*7*, *62*). The variants included were rare in the population with mean allele frequency <1% in dbSNP147 and with only 0-1 homozygotes in the adult genome database gnomAD. Additionally, variants were non-synonymous and/or located within splice-sites. Subsequently, variant severity were stratified based on protein impact (truncating frameshift or nonsense variants, essential or extended splice-site variants, and missense variants). Splice-site variants were assessed by *in silico* tools MaxEnt, NNSPLICE, SpliceSite Finder, and GeneSplicer splice-site variant prediction scores as well as conservation across human splice-sites as described previously (*7*, *62*). Missense variants were assessed based on SIFT, MutationTaster, PolyPhen 2.0 and CADD conservation prediction scores and evolutionary conservation based on manually derived multiple sequence alignments.

### ClinVar Database Query

The ClinVar Database (*32*) was queried for variants in *NOS1AP* and *C1orf226* that were not large structural variants. These variants were analyzed as above for rare population prevalence (<1% allele frequency) and predicted deleterious effects on protein coding (truncating variant or missense variant with 2+ strong *in silico* prediction scores) and/or splicing (2+ strong *in silico* prediction scores). Variant submissions that were predicted to be deleterious were further evaluated by contacting the designated submitter for additional genetic information (zygosity, method of detection and other detected variants) and clinical information. The ClinVar subject had CLIA exome sequencing performed (3Billion, South Korea) with confirmed enrichment for known NS-associated disease genes.

### Mouse breeding and maintenance for *Nos1ap^Ex3-/Ex3-^* mice

The animal experimental protocols were reviewed and approved by the Institutional Animal Care and Use Committee at the Boston Children’s Hospital (#18-12-3826R). All mice were handled in accordance with the Guidelines for the Care and Use of Laboratory Animals. Mice were housed under pathogen-free conditions with a light period from 7:00 AM to 7:00 PM and had *ad libitum* access to water and rodent chow. *Nos1ap^Ex3-/Ex3-^* mice were a kind gift from Dr. Norihiro Kato from National Center for Global Health and Medicine in Japan. In brief, a linearized vector with targeted deletion of *Nos1ap* Exon 3 was electroporated into ES cells, positive clones selected by G418 on embryonic fibroblast feeder cells and targeted ES cells then microinjected into C57BL/6 blastocysts. Resulting male chimeras were backcrossed onto C57BL/6 mice and heterozygous Nos1ap^Ex3-/^**^+^** mice N4 generations were intercrossed to generate homozygous Nos1ap^Ex3-/Ex3-^ mice. Genotyping was performed by multiplex PCR using the following primers: #1:CTTTGTCTTCTGCTTCGCC, #2:ACACTACCATTTGGTCTCC, #3:TCAAGACCGACCTGTCC and #4:CAATAGCAGCCAGTCCC.

### Mouse generation and maintenance of *Nos1ap^Ex4-/E4-^* and *Gm7694^-/-^*

The animal experimental protocols were reviewed and approved by (Dalhousie Animal Care Protocol #21-120 and #20-116). All mice were handled in accordance with the Canadian Council on Animal Care (CCAC).

*Nos1ap^Ex4-/E4-^* were generated as follows. Mice were ordered and generated at Baylor using the KOMP Tm1a heterozygous knockout mice for gene *Nos1ap* (MGI ID 1917979, Knockout-First - Reporter Tagged Insertion (Promotor Driven Cassette)). The Knockout-First Reporter Tagged mice NOS1AP^+/tm1a^ mice were then crossed with CMV-Cre mice from Jax labs (B6.CTg(CMV- Cre)1 Cgn/J (Stock #006054)) to yield animals with complete excision of the neo cassette and exon 4 but a residual *LacZ* reporter gene in the locus (*Nos1ap^+/Ex4-^*). PCR amplification of isolated genomic DNA is used for genotyping using the following primers to detect the mutant allele (*Nos1ap^Ex4-^*) 5’-CGGATAAACGGAACTGGAAA-3’ and 5’-TAATCACGACGCGCTGTATC-3’. For the wild type allele the following primer sets are used 5’-CAATTCATGGCAAGCAAAAC-3’ and 5’- ATTTCTCTTCCTCCGCACCT-3’. See Genbank file https://www.i-dcc.org/imits/targ_rep/alleles/26526/escell-clone-genbank-file. Mice were maintained on a C57BL/6 background. They were in-crossed to generate wildtype, heterozygous and homozygous mice.

*Gm7694^-/-^* mice were generated by the Center for Mouse Genome Modification, University of Connecticut Health Center. LoxP sites were placed around exons 1 and 2 of the *Gm7694* gene that undergoes intergenic splicing with *Nos1ap*. Mouse embryonic stem cells targeting *Gm7694* were generated by blastocyst injection and tested for germline transmission. The resulting mosaic mice were crossed with ROSA26-Flpe mice to remove an internal neo cassette. To generate a deleted allele (*Gm7694^-^*), the LoxP containing mice were crossed with the CMV-Cre mice from Jackson. Loss of *Gm7694* exons 1 and 2 were confirmed by PCR. The KO allele genotyping is done with the following primers 5’-GCCCCTCCTAATTCCAAGTG-3’ and 5’- ACTGACCCAGCAAACCAACT-3’ and the wild type allele is detected with the following primers 5’-ATACGGGCCCTCTCTTAAC-3’ and 5’-ACTGACCCAGCAAACCAACT-3’. *Gm7694^-/+^* mice were in-crossed to generate wildtype, heterozygous and homozygous mice. Mice were maintained on a C57BL/6 background.

### Urine analysis

Urine was collected by placing mice in collection cages with *ad libitum* access to water overnight (16 h). Upon collection, samples were immediately frozen and stored at -80 °C and only thawed on ice prior to urine albumin and creatinine measurements once. Urinary albumin levels were determined using the Albumin Blue Fluorescent Assay Kit (Active Motif) in combinations with standard dilutions prepared from mouse serum albumin (Equitech Bio Inc.). Urine creatinine was measured using QuantiChrom™ Creatinine Assay Kit (BioAssay Systems). In-gel protein detection was performed with Coomassie blue dye.

### Whole blood analysis

200 μl of blood were collected once a month via facial vein bleeding method and collected in lithium heparin tubes. Blood samples were then immediately analyzed with the Vetscan_®_ VS2 Chemistry Analyzer using Comprehensive Diagnostic Profile rotors.

### Histological analysis

The kidney tissues were fixed in 4% paraformaldehyde (PFA), sectioned (5 μm thickness) and stained with periodic acid-Schiff (PAS) following standard protocols for histological examination. For mesangial matrix deposition, 25 images (at 40X magnification) per animal were obtained to detect glomerular tufts and stalk. Quantitative analysis of PAS-stained sections was performed in a semiautomated manner using an ImageJ macro. Briefly, glomeruli were outlined manually and cropped. Color splitting was performed to yield images of component colors that reflected the PAS-positive extracellular matrix and nuclei or nuclei alone. Thresholding and particle analysis were performed to determine the percent area in these channels to calculate the percent glomerular area comprised of PAS-positive matrix. For tubular area, 5 images (at 20X magnification) of the corticomedullary junction distributed across a longitudinal section of a kidney section were obtained for each animal. Quantitative analysis of the tubular lumen area was determined using a semiautomated ImageJ macro. Thresholding was performed on images to detect open tubular lumen, and particular analysis was performed to calculate the percentage of positive area.

### Ultrastructural analysis

Kidney tissues were perfuse-fixed with 2.5% glutaraldehyde, 1.25% PFA in 0.1 M sodium cacodylate buffer (pH 7.4) and then fixed in 2.5% glutaraldehyde, 1.25% PFA, and 0.03% picric acid in 0.1 M sodium cacodylate buffer (pH 7.4) overnight at 4 °C. Samples were then washed with 0.1 M phosphate buffer, post-fixed with 1% OsO_4_ dissolved in 0.1 M phosphate-buffered saline (PBS) for 2 h, dehydrated in ascending gradual series (50‒100%) of ethanol, and infiltrated with propylene oxide. Samples were embedded using the Poly/Bed 812 kit (Polysciences) according to manufacturer’s instructions. After pure fresh resin embedding and polymerization in a 65°C oven (TD-700, DOSAKA, Japan) for 24 h, sections of approximately 200–250 nm thickness were cut and stained with toluidine blue for light microscopy. Sections of 70-nm thickness were double stained with 6% uranyl acetate (EMS, 22400) for 20 min and lead citrate (Fisher) for 10 min for contrast staining. The sections were cut using Reichert Ultracut-S/LEICA EM UC-7 (Leica) with a diamond knife (Diatome) and transferred on to copper and nickel grids. Sections were evaluated by transmission electron microscopy (JEOL 1200EX) at an acceleration voltage of 80 kV. All steps, including image acquisition, were performed in a blinded manner by independent persons. For each animal, ∼21 images of 3-4 glomeruli were acquired for assessment of podocyte foot process density and glomerular basement membrane thickness only perpendicularly cut capillary loops evaluated.

### Mouse Treatment Studies

For lisinopril studies, mice were treated with 100 or 200 mg/L of lisinopril (Sigma, in their drinking water as previously described(*63*). Lisinopril or vehicle (water) containing drinking water was changed three times weekly. Urine samples were collected every two weeks. Blood samples were collected at an early and late timepoint in the study. Mice were monitored every 1-2 days for survival for >20 weeks of treatment.

For dynamin activating agent Bis-T-23 (Aberjona Labs), 20 or 30 mg/kg were administered daily to mice by intraperitoneal injection as previously described (*46*). First, urine was collected daily from four-week-old FVB/N-*Nos1ap^Ex3-/Ex3-^* mice receiving Bis-T-23 for seven days to detect reduction in albuminuria prior to severe disease progression. In a second study, homozygotes were treated daily, and urine samples were collected immediately prior to injection and hourly for the subsequent three hours to detect transient changes in albuminuria.

### RT-PCR Studies

C57BL/6 mice kidneys were isolated from 8-week-old male mice. RNA isolation was performed using RNAeasy Mini Kit (Qiagen). Total RNA was converted into cDNA using the TeloPrime Full- Length cDNA Amplification Kit V2 (Lexogen). cDNA was amplified using HotStarTaq DNA Polymerase and the following primers: Ex7_9_F, CGAGGTGTGACTGATCTGGA; Ex7_GEx2_F, GTGTGACTGATCTGGATGCC; Ex7_GEx2_R, GTGTTCTTGTGTATTAGGTCCGG; Ex8_Gex2_F, TCCACTCACCACCAGATGC; Ex8_Gex2_R, CCTCCTGGGTGTTCTTGTGT; Ex9_GEx2_R, TAGATTGCTTAGGACGGGCT; Ex2-3_Gex_F:, CGCAGAATCCGGTATGAGTT; Ex3_Gex_F, TCTCTGTGGACGGTGTCAAG; Ex4_Gex_F, CTGGTGATGCAGGACCCTAT; Ex7- 9-F_Ex10UTR-R1, ACCCTGGGTGTTGTTCTCAG; Ex8-10-F_Ex10UTR-R2, ACCACCTTCACCAGCTCCTC. Human kidney total RNA was purchased from Takara. The sample was isolated from a post-mortem kidney sample from an adult female who died from sudden death unrelated to kidney disease. RNA was reverse transcribed using the iScript cDNA Synthesis Kit (Bio-Rad). cDNA was amplified using HotStarTaq DNA Polymerase and the following primers: NEX1UTRtoATG_F: CGGGTAACCATGCCTAGCAAAAC; FL_NOS1AP_R2: ACCTACACGGCGATCTCATCATC; FL_FUSION_NOS_C1ORF_R1: CTATTCAAAGGACAGCAGGTCTG. PCR products were analyzed by agarose gel electrophoresis and subsequent Sanger sequencing.

### Mouse Bulk RNA sequencing (RNAseq) re-analysis

50 bp single end reads sequenced from wildtype C57BL/6J mice kidney samples at age 0, 2, 4, 8 and 79 weeks were downloaded from Gene Expression Omnibus (GEO) dataset GSE225622 (*64*). 101 bp paired end reads sequenced from wildtype C57BL/6J mouse kidney samples at 8 weeks of life were downloaded from dataset GSE145053 (*65*). All samples for each time point were pooled into a single dataset (**Supplementary Table 2**). We used trimmomatic v0.39 to trim the low-quality next generation sequencing (NGS) reads (-threads 20 ILLUMINACLIP:TruSeq3- PE.fa:2:30:10 LEADING:3 TRAILING:3 SLIDINGWINDOW:4:15 MINLEN:36) (*66*). Subsequently, only the high-quality trimmed reads were aligned to the mouse reference genome using STAR v2.7.2b (*67*). All aligned reads spanning the splice site coordinates (GRCm38/mm10) chr1:170,318,738 on Nos1ap gene and chr1:170,302,838 on Gm7694 gene within (i) a 5 bp window with minimum 1 bp overlap on either side of the splice junction or (ii) fixed 10 bp window (5 bp on both sides of junction) were counted in IGV v2.15.2 (*68*). Reads that did not show spliced alignment but spanned the splice junction on Nos1ap were counted as canonical reads while reads that show spliced alignment were counted as intergenic reads. A ratio of intergenic to total reads was computed where sum of all reads spanning the Nos1ap splice junction were taken as the total.

### Bulk Long Read RNAseq re-analyses from Human Tissues (ENCODE)

To verify that the intergenic transcript is expressed in human tissues, we interrogated long read sequencing from the ENCODE project (*69*). We downloaded 31 bam files from 1 kidney, 3 colon, 10 brain and 17 heart adult tissues sequenced using Pacific Biosciences Sequel technology (sample details in supplement table 2). We visually inspected each bam file using the IGV browser (*70*). We, first, identified “long transcripts” which began before or within exon 3 of *NOS1AP* (the earliest exon where we found causative disease variants in human or mice). We, then, quantified canonical transcripts, reads that extend into the UTR of *NOS1A*P exon 10, and intergenic transcripts that extend into the second coding exon of C1orf226. To visualize the long-read sequencing, we adapted the output from the alignment track in the IGV browser. For representative examples, we selected the bam file from each tissue with the most intergenic reads. For clarity of the figure and to emphasize the long reads, we filtered to reads aligned to the positive strand and hid base-pair mismatches.

### Bulk Short Read RNAseq re-analyses from Human Kidney Tissue (NEPTUNE)

To quantify the intergenic transcripts in human kidney tissues, we utilized micro-dissected glomerular and tubulointerstitial compartments from the NEPTUNE cohort, which included patients with nephrotic syndrome as well as healthy tissue from tumor nephrectomies and living donors (*71*). Biopsies were prepared using the Clontech SMARTSeq v4 kit and sequenced using Illumina HiSeq 2500, resulting in 150bp unstranded, paired-end reads. For each sample, individual reads were checked for adapter content, GC content, and per-base sequence quality using FastQC. FastQ Screen was used to detect contamination of foreign or disproportionate amounts of off-target RNA (*72*). Sequenced reads were then aligned to the human reference genome GRCh38 using STAR 2.5.2b and mapped reads were inspected for distribution across introns, exons, UTR, and intergenic regions using picardtools (*67*) (http://broadinstitute.github.io/picard). We then used SAMtools to filter out duplicates, keep only properly paired reads, and filter out reads with low mapping quality (<20)(*73*). We filtered each bam file to the start of *NOS1AP* and end of *C1orf22* with a 1kb flank (CRCh37/hg19 chr1:162,024,660-162,360,673). The 150 bp reads were analyzed in exon 10 of *NOS1AP* that either splice to *C1orf266* or extend through exon 10. To do this, we defined a 10 bp sequence for each the canonical (CTTAGGTAGG) and intergenic (CTTAGTTGAC) transcripts, which ensured at least 5 matched nucleotides before and after the splice site. For each bam file, we tabulated both sequences. Reads that did not completely span either sequence were excluded.

### Mouse kidney podocytes deep proteomics re-analysis

To assess if peptides corresponding to either canonical or intergenic Nos1ap isoforms are abundant in mouse glomerular podocytes, deep proteomics data from Rinschen et al (*29*) was re- analyzed. In brief, liquid chromatography-tandem mass spectrometry was performed on FACS-sorted podocytes and other glomerular cells from isolated mouse glomeruli. Cells were digested using trypsin and, to increase peptide detection, six-layered SCX resin and fractionation using six different buffers was performed. Proteomics raw data was downloaded from PRIDE (PXD003306). Next, data was re-analyzed with MaxQuant (v.1.6.10) (*74*) using the Uniprot SwissProt Mouse database (downloaded in April 2021) amended by the sequences of non-canonical intergenic Nos1ap, Gm7694 and the unique C-terminus of the canonical Nos1ap. In total, 23 peptides were identified.

### Nos1ap Immunoprecipitation from mouse fibroblasts

For immunoprecipitation assays, fibroblasts were lysed in NP40 lysis buffer (10% glycerol, 1% NP-40, 137mM NaCL, 20mM Tris [pH 8.0] containing 1mM phenylmethylsulfonyl fluoride (PMSF), 10ug/ml aprotinin, and 10 ug/ml leupeptin. Clarified neonatal kidney lysates were then precipitated with an antibody raised against a region of canonical rat Nos1ap (amino acids 304-503) that is partially shared by intergenic Nos1ap (amino acids 304-443) (Antibody R300, Santa Cruz sc- 9138) (*23*, *26*). Clarified mouse embryonic fibroblasts were precipitated with an antibody raised against a region of canonical rat Nos1ap (amino acids 366-503) that is partially shared by intergenic Nos1ap (amino acids 366-443) (Antibody 2093) (*23*, *26*). Following an overnight incubation at 4 C, Protein A Sepharose was added and incubated for 1 hour at 4 C. The precipitated proteins were then washed 3 times in NP40 lysis buffer before the addition of 2 x Sample Buffer. Protein samples were then resolved on 8% or 10% SDS-PAGE gels, transferred to PVDF membrane (EMD, Millipore, Billerica, MA) and then blocked in 5% skim milk in 1x TBST for 1 hour. Membranes were then incubated in primary antibodies overnight at 4 C: R300 (1:1000) for neonatal kidney; Antibody 2093 (1:1000) for mouse fibroblasts; or GSTLong (1:5000) (raised against an immunogen containing intergenic Nos1ap amino acids 446-715) (*23*, *26*). Blots were then washed 3 times in TBST then incubated in TBST containing HRP-conjugated secondary at 1:10,000 for 1 hour at RT. Membranes were then washed 3 times in TBST prior to signal detection using ECL reagent (BioRad). Blots were imaged using a ChemiDoc (BioRa).

### Statistics

Graphpad Prism 8.0.0 software was used to perform statistical testing between groups.

### Web Resources

UCSC Genome Browser, genome.ucsc.edu Ensembl Genome Browser, www.ensembl.org gnomAD browser 2.0.3., gnomad.broadinstitute.org

## Supporting information

Supplement complete

## Data Availability

All data produced in the present study are either already contained in the manuscript or remaining data available upon reasonable request to the authors.

## Author Contributions

1. F.B., D.S., L.L., V.G., K.K., M.Q., V.S., A.R., D.B., K.L., K.S., L.M.M., S.S., J.F., and A.J.M. designed and/or performed animal model studies.
2. F.B., B.I., L.S., S.G.C, A.C.G., M.T.M., M.G.S., and A.J.M. performed transcriptomic and proteomic analyses.
3. F.B., V.G., A.R., K.L., M.H.A., M.M.S., M.S., J.K., F.H. and A.J.M recruited patients, gathered detailed clinical information, and performed genetic analysis.
4. All authors critically reviewed the paper.
5. A.J.M, F.B., and F.H. conceived of and directed the project.
6. A.J.M, F.B., and F.H. prepared the manuscript.

## Data Sharing Statement

All data needed to evaluate the conclusions in the paper are present in the paper and/or the Supplementary Materials.

