## Supplement complete for "Recessive variants in the intergenic *NOS1AP-C1orf226* locus cause monogenic kidney disease responsive to anti-proteinuric treatment"

**Supplementary Document 1. Members of the Nephrotic Syndrome Study Network (NEPTUNE)**

**NEPTUNE Collaborating Sites**

*Atrium Health Levine Children's Hospital, Charlotte, SC*: Susan Massengill[*], Layla Lo[#]
*Cleveland Clinic, Cleveland, OH*: Katherine Dell[*], John O'Toole[*], John Sedor[**], Victoria Grange[#]
*Children's Hospital, Los Angeles, CA*: Ian Macumber[*], Alyssa Parry[#]
*Children's Mercy Hospital, Kansas City, MO*: Tarak Srivastava[*], Kelsey Markus[#]
*Cohen Children's Hospital, New Hyde Park, NY*: Christine Sethna[*], Suzanne Vento[#]
*Columbia University, New York, NY:* Pietro Canetta[*]
*Duke University Medical Center, Durham, NC:* Opeyemi Olabisi[*], Rasheed Gbadegesin[**], Maurice Smith[#]
*Emory University, Atlanta, GA:* Laurence Greenbaum[*], Chia-shi Wang[*], Emily Yun[#]
*The Lundquist Institute, Torrance, CA:* Sharon Adler[*], Janine LaPage[#]
*John H Stroger Cook County Hospital, Chicago, IL:* Amatur Amarah[*]
*Johns Hopkins Medicine, Baltimore, MD:* Meredith Atkinson[*], Sara Boynton[#]
*Mayo Clinic, Rochester, MN:* John Lieske, Marie Hogan, Fernando Fervenza
*Medical University of South Carolina, Charleston, SC:* David Selewski[*], Cheryl Alston[#]
*Montefiore Medical Center, Bronx, NY:* Kim Reidy[*], Michael Ross[*], Frederick Kaskel[**], Patricia Flynn[#]
*New York University Medical Center, New York, NY:* Laura Malaga-Dieguez[*], Olga Zhdanova[**], Laura Jane Pehrson[#], Melanie Miranda[#]
*The Ohio State University College of Medicine, Columbus, OH*: Salem Almaani[*], Laci Roberts[#]
*Stanford University, Stanford, CA:* Richard Lafayette[*], Shiktij Dave[#]
*Temple University, Philadelphia, PA:* Iris Lee[**]
*Texas Children's Hospital at Baylor College of Medicine, Houston, TX*: Shweta Shah[*], Sadaf Batla[# #]
*University Health Network Toronto:* Heather Reich[*], Michelle Hladunewich[**], Paul Ling[#], Martin Romano[#]
*University of California at San Francisco, San Francisco, CA*: Paul Brakeman[*], Daniel Schrader
*University of Colorado Anschutz Medical Campus, Aurora, CO*: James Dylewski[*] Nathan Rogers[#]
*University of Kansas Medical Center, Kansas City, KS*: Ellen McCarthy[*], Catherine Creed[#]
*University of Miami, Miami, FL:* Alessia Fornoni[*], Miguel Bandes[#]
*University of Michigan, Ann Arbor, MI:* Matthias Kretzler[*], Laura Mariani[*], Zubin Modi[*], A Williams[#], Roxy Ni[#]
*University of Minnesota, Minneapolis, MN:* Patrick Nachman[*], Michelle Rheault[*], Amy Hanson[#], Nicolas Rauwolf[#]
*University of North Carolina, Chapel Hill, NC:* Vimal Derebail[*], Keisha Gibson[*], Anne Froment[#], Mary Mac McGown Collie[#]
*University of Pennsylvania, Philadelphia, PA:* Lawrence Holzman[*], Kevin Meyers[**], Krishna Kallem[#], Aliya Edwards[#]
*University of Texas San Antonio, San Antonio, TX*: Samin Sharma[**]
*University of Texas Southwestern, Dallas, TX:* Elizabeth Roehm[*], Kamalanathan Sambandam[**], Elizabeth Brown[**], Jamie Hellewege
*University of Washington, Seattle, WA:* Ashley Jefferson[*], Sangeeta Hingorani[**], Katherine Tuttle[**§], Linda Manahan [#], Emily Pao[#], Kelli Kuykendall[§]
*Wake Forest University Baptist Health, Winston-Salem, NC:* Jen Jar Lin[**]
*Washington University in St. Louis, St. Louis, MO*: Vikas Dharnidharka[*]

**Data Analysis and Coordinating Center:** *University of Michigan:* Matthias Kretzler[*], Brenda Gillespie[**], Laura Mariani[**], Zubin Modi[**], Eloise Salmon[**], Howard Trachtman[**], Tina Mainieri, Gabrielle Alter, Michael Arbit, Hailey Desmond, Sean Eddy, Damian Fermin, Wenjun Ju, Maria Larkina, Chrysta Lienczewski, Rebecca Scherr, Jonathan Troost, Amanda Williams, Yan Zhai; *Arbor Collaborative for Health:* Colleen Kincaid, Shengqian Li, Shannon Li; *Cleveland Clinic:* Crystal Gadegbeku[**], *Duke University:* Laura Barisoni[**]; John Sedor[**], *Harvard University:* Matthew G Sampson[**]; *Northwestern University:* Abigail Smith[**]; *University of Pennsylvania:* Lawrence Holzman[**], Jarcy Zee[**]

**Digital Pathology Committee:** Carmen Avila-Casado *(University Health Network)*, Serena Bagnasco *(Johns Hopkins University)*, Lihong Bu *(Mayo Clinic)*, Shelley Caltharp *(Emory University)*, Clarissa Cassol *(Arkana)*, Dawit Demeke *(University of Michigan)*, Brenda Gillespie *(University of Michigan)*, Jared Hassler *(Temple University)*, Leal Herlitz *(Cleveland Clinic)*, Stephen Hewitt *(National Cancer Institute)*, Jeff Hodgin *(University of Michigan)*, Danni Holanda *(Arkana)*, Neeraja Kambham *(Stanford University)*, Kevin Lemley, Laura Mariani *(University of Michigan)*, Nidia Messias

*(Washington University)*, Alexei Mikhailov *(Wake Forest)*, Vanessa Moreno *(University of North Carolina)*, Behzad Najafian *(University of Washington)*, Matthew Palmer *(University of Pennsylvania)*, Avi Rosenberg *(Johns Hopkins University)*, Virginie Royal *(University of Montreal)*, Miroslav Sekulik *(Columbia University)*, Barry Stokes *(Columbia University)*, David Thomas *(Duke University)*, Ming Wu *(University of New York)*, Michifumi Yamashita *(Cedar Sinai)*, Hong Yin *(Emory University)*, Jarcy Zee *(University of Pennsylvania)*, Yiqin Zuo *(University of Miami)*. Co-Chairs: Laura Barisoni *(Duke University)*, Cynthia Nast *(Cedar Sinai)*.

**Supplementary Document 2. Bottom-up proteomics analysis of canonical and non-canonical Nos1ap from mouse glomeruli**

Proteomics raw data from Project PXD003306 “Deep mapping of the mouse podocyte proteome”, corresponding to FACS-sorted podocytes and non-podocyte cells of the glomerulus (Rinschen MM et al. 2018, PMID:29791858) were re-analysed in MaxQuant (v.1.6.10) using the Uniprot SwissProt Mouse database (downloaded in April 2021) amended by the sequences of non-canonical intergenic Nos1ap, Gm7694 and the unique C-terminus of the canonical Nos1ap. In total, 23 peptides were identified. Nineteen of which are shared between the canonical and the non-canonical Nos1ap, and four localized at the unique C-terminal region of the non-canonical Nos1ap.

**Nos1ap peptides identified in mouse kidneys**

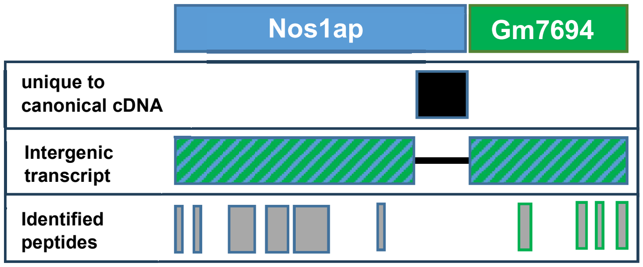


No peptides aligning to the unique C-terminal part of the canonical Nos1ap (black rectangle in schematic overview) were detected despite theoretical trypsin cleavage sites, yielding a least five potentially detectable peptides with lengths ranging from 7 to 25 amino acids.

```
                  Tryps                     Tryps
Tryps Tryps         |                 Tryps |              Tryps

   |     |     |                          | |                  |
   GRHDCLVKLECFRFLPPEDTQPMMAQGEPLLGGLELIKFRESGIASEYESNTDESEERDS
1  ---------+---------+---------+---------+---------+---------+  60


     Tryps  Tryps

         |      |
   WSQEELPRLLNVLQRQELGDSLDDEIAV
61 ---------+---------+--------  88
```

| Position of cleavage site | Cleaving enzyme | Resulting peptide sequence | Peptide length [aa] | Peptide Mass [Da] |
|---|---|---|---|---|
| 2 | Trypsin | GR | 2 | 213.255 |
| 8 | Trypsin | HDCLVK | 6 | 713.850 |
| 13 | Trypsin | LECFR | 5 | 666.793 |
| 38 | Trypsin | FLPPEDTQPMMAQGEPLLGGLELIK | 25 | 2725.210 |
| 40 | Trypsin | FR | 2 | 321.379 |
| 58 | Trypsin | ESGIASEYESNTDESEER | 18 | 2031.972 |
| 68 | Trypsin | DSWSQEELPR | 10 | 1246.299 |
| 75 | Trypsin | LLNVLQR | 7 | 855.048 |
| 88 | End of sequence | QELGDSLDDEIAV | 13 | 1403.462 |

*Theoretical cleavage sites and peptides derived from the C-terminus of canonical mouse Nos1ap. Potentially detectable peptides are highlighted.*

No peptides were detected spanning the intergenic region of the non-canonical Nos1ap. An *in-silico* digestion of the non-canonical Nos1ap with trypsin yielded no detectable peptides from the intergenic region. Of note, the resulting peptide would exceed the standard peptide lengths for identification.

| Position of cleavage site | Cleaving enzyme | Resulting peptide sequence | Peptide length [aa] | Peptide Mass [Da] |
|---|---|---|---|---|
| 431 | Trypsin | SGALPVLCESTTPKPEDLHSPLLGAGLADFAHPAGSPLVDHSMFENLNTTLTPK | 54 | 5596.326 |

*Theoretical tryptic peptide spanning the intergenic region of non-canonical mouse Nos1ap. Intergenic slice junction in red.*

**Supplementary Table 1. RNAseq Datasets of Mouse Kidney Samples for Re-analysis**

| Publication | GSE Accession Number | Sample | FilesNames |
|---|---|---|---|
| PMID: 37205355<br><br>47 samples 107 files | GSE225622 (Wk0) 8 samples 8 files | GSM7051385 31_F1_P0_C57BL/6J<br>GSM7051386 32_F2_P0_C57BL/6J<br>GSM7051387 73_fp0_3_C57BL/6J<br>GSM7051388 74_fp0_3_C57BL/6J<br>GSM7051389 36_M4_P0_C57BL/6J<br>GSM7051390 33_M1_P0_C57BL/6J<br>GSM7051391 34_M2_P0_C57BL/6J<br>GSM7051392 35_M3_P0_C57BL/6J | SRR23555586<br>SRR23555585<br>SRR23555584<br>SRR23555583<br>SRR23555582<br>SRR23555581<br>SRR23555580<br>SRR23555579 |
| | GSE225622 (Wk2) 10 samples 30 files | GSM7051393 21_F1_2wk_C57BL/6J<br>GSM7051394 22_F2_2wk_C57BL/6J<br>GSM7051395 23_F3_2wk_C57BL/6J<br>GSM7051396 24_F4_2wk_C57BL/6J<br>GSM7051397 26_F5_2wk_C57BL/6J<br>GSM7051398 25_M1_2wk_C57BL/6J<br>GSM7051399 27_M2_2wk_C57BL/6J<br>GSM7051400 28_M3_2wk_C57BL/6J<br>GSM7051401 29_M4_2wk_C57BL/6J<br>GSM7051402 30_M5_2wk_C57BL/6J | SRR23555578<br>SRR23555577<br>SRR23555576<br>SRR23555518<br>SRR23555519<br>SRR23555520<br>SRR23555515<br>SRR23555516<br>SRR23555517<br>SRR23555512<br>SRR23555513<br>SRR23555514<br>SRR23555509<br>SRR23555510<br>SRR23555511<br>SRR23555506<br>SRR23555507<br>SRR23555508<br>SRR23555503<br>SRR23555504<br>SRR23555505<br>SRR23555500<br>SRR23555501 |

| | | | |
|---|---|---|---|
| | | | SRR23555502<br>SRR23555497<br>SRR23555498<br>SRR23555499<br>SRR23555494<br>SRR23555495<br>SRR23555496 |
| | GSE225622 (Wk4)<br>10 samples 30 files | GSM7051403 16_F1_4wk_C57BL/6J<br>GSM7051404 17_F2_4wk_C57BL/6J<br>GSM7051405 18_F3_4wk_C57BL/6J<br>GSM7051406 19_F4_4wk_C57BL/6J<br>GSM7051407 20_F5_4wk_C57BL/6J<br>GSM7051408 11_M1_4wk_C57BL/6J<br>GSM7051409 12_M2_4wk_C57BL/6J<br>GSM7051410 13_M3_4wk_C57BL/6J<br>GSM7051411 14_M4_4wk_C57BL/6J<br>GSM7051412 15_M5_4wk_C57BL/6J | SRR23555493<br>SRR23555492<br>SRR23555491<br>SRR23555490<br>SRR23555489<br>SRR23555488<br>SRR23555487<br>SRR23555486<br>SRR23555485<br>SRR23555484<br>SRR23555483<br>SRR23555482<br>SRR23555481<br>SRR23555480<br>SRR23555479<br>SRR23555478<br>SRR23555477<br>SRR23555476<br>SRR23555475<br>SRR23555474<br>SRR23555473<br>SRR23555575<br>SRR23555574<br>SRR23555573<br>SRR23555472<br>SRR23555471<br>SRR23555470<br>SRR23555469 |

| | | | |
|---|---|---|---|
| | | | SRR23555468<br>SRR23555467 |
| | GSE225622 (Wk8)<br>10 samples 30 files | GSM7051413 6_F1_8wk_C57BL/6J<br>GSM7051414 7_F2_8wk_C57BL/6J<br>GSM7051415 8_F3_8wk_C57BL/6J<br>GSM7051416 9_F4_8wk_C57BL/6J<br>GSM7051417 10_F5_8wk_C57BL/6J<br>GSM7051418 1_M1_8wk_C57BL/6J<br>GSM7051419 2_M2_8wk_C57BL/6J<br>GSM7051420 3_M3_8wk_C57BL/6J<br>GSM7051421 4_M4_8wk_C57BL/6J<br>GSM7051422 5_M5_8wk_C57BL/6J | SRR23555564<br>SRR23555565<br>SRR23555566<br>SRR23555561<br>SRR23555562<br>SRR23555563<br>SRR23555558<br>SRR23555559<br>SRR23555560<br>SRR23555557<br>SRR23555556<br>SRR23555555<br>SRR23555554<br>SRR23555553<br>SRR23555552<br>SRR23555470<br>SRR23555471<br>SRR23555472<br>SRR23555467<br>SRR23555468<br>SRR23555469<br>SRR23555464<br>SRR23555465<br>SRR23555466<br>SRR23555461<br>SRR23555462<br>SRR23555463<br>SRR23555458<br>SRR23555459<br>SRR23555460 |
| | GSE225622 (Wk79)<br>9 samples. 9 files | GSM7051443 75_vf1.5yr_1_C57BL/6NCrl<br>GSM7051444 76_vf1.5yr_2_C57BL/6NCrl | SRR23555393<br>SRR23555392 |

| | | | |
|---|---|---|---|
| | | GSM7051445 77_vf1.5yr_3_C57BL/6NCrl<br>GSM7051446 78_vf1.5yr_4_C57BL/6NCrl<br>GSM7051447 79_vf1.5yr_5_C57BL/6NCrl<br>GSM7051448 80_vf1.5yr_2_C57BL/6NCrl<br>GSM7051449 81_vf1.5yr_3_C57BL/6NCrl<br>GSM7051450 82_vf1.5yr_4_C57BL/6NCrl<br>GSM7051451 83_vf1.5yr_5_C57BL/6NCrl | SRR23555391<br>SRR23555390<br>SRR23555389<br>SRR23555388<br>SRR23555387<br>SRR23555386<br>SRR23555385 |
| PMID: 32978267<br>GEO: GSE145053<br>4 samples 12 files | GSM4305441 | 8 week old C57BL/6J #1 | SRR11058622<br>SRR11058623<br>SRR11058624 |
| | GSM4305442 | 8 week old C57BL/6J #2 | SRR11058625<br>SRR11058626<br>SRR11058627 |
| | GSM4305443 | 8 week old C57BL/6J #3 | SRR11058628<br>SRR11058629<br>SRR11058630 |
| | GSM4305444 | 8 week old C57BL/6J #4 | SRR11058631<br>SRR11058632<br>SRR11058633 |

**Supplementary Table 2. Bulk Long Read RNAseq Summary Data from ENCODE**

| Organ | Assession # | Total long counts (*NOS1AP* Exon 1-3 to either *NOS1AP* 3' UTR or into *C1orf226* Exon 2) | Intergenic Counts (*NOS1AP* Exon 1-3 to into *C1orf226* Exon 2) | Canonical Ratio | Intergenic Ratio | Age | Sex | Tissue |
|---|---|---|---|---|---|---|---|---|
| Kidney | ENCFF239CWJ | 3 | 1 | 66.67 | 33.33 | 47 | female | kidney tissue |
| Brain | ENCFF678TEN | 7 | 1 | 85.71 | 14.29 | 90+ | female | dorsolateral prefrontal cortex tissue with Alzheimer's disease |
| Brain | ENCFF114VSO | 8 | 0 | 100.00 | 0.00 | 90+ | female | dorsolateral prefrontal cortex tissue |
| Brain | ENCFF877QJZ | 10 | 0 | 100.00 | 0.00 | 90+ | male | dorsolateral prefrontal cortex tissue with Alzheimer's disease |
| Brain | ENCFF378STM | 8 | 0 | 100.00 | 0.00 | 90+ | female | dorsolateral prefrontal cortex tissue with Alzheimer's disease |
| Brain | ENCFF564ONS | 9 | 1 | 88.89 | 11.11 | 90+ | female | dorsolateral prefrontal cortex tissue with Alzheimer's disease |
| Brain | ENCFF406GQU | 17 | 1 | 94.12 | 5.88 | 90+ | female | dorsolateral prefrontal cortex tissue with Alzheimer's disease |
| Brain | ENCFF222UTL | 12 | 0 | 100.00 | 0.00 | 79 | female | dorsolateral prefrontal cortex tissue |
| Brain | ENCFF279ABL | 3 | 0 | 100.00 | 0.00 | 88 | female | dorsolateral prefrontal cortex tissue |
| Brain | ENCFF319FBW | 1 | 0 | 100.00 | 0.00 | 53 | male | astrocyte in vitro differentiated cells |
| Heart | ENCFF626QRV | 1 | 0 | 100.00 | 0.00 | 41 | female | cardiac septum tissue |
| Heart | ENCFF613SDS | 2 | 2 | 0.00 | 100.00 | 46 | female | right cardiac atrium tissue |
| Heart | ENCFF291EKY | 2 | 1 | 50.00 | 50.00 | 60 | male | right cardiac atrium tissue |
| Heart | ENCFF173JOL | 3 | 3 | 0.00 | 100.00 | 60 | male | right ventricle myocardium inferior tissue |
| Heart | ENCFF840OVC | 5 | 2 | 60.00 | 40.00 | 60 | male | right ventricle myocardium superior tissue |
| Heart | ENCFF132YCF | 8 | 5 | 37.50 | 62.50 | 60 | male | left ventricle myocardium inferior tissue |
| Heart | ENCFF850YMO | 2 | 1 | 50.00 | 50.00 | 60 | male | left ventricle myocardium superior tissue |
| Heart | ENCFF623IBV | 3 | 1 | 66.67 | 33.33 | 40 | male | heart left ventricle tissue |
| Heart | ENCFF018PZX | 2 | 0 | 100.00 | 0.00 | 40 | male | heart right ventricle tissue |
| Heart | ENCFF502LAB | 2 | 1 | 50.00 | 50.00 | 46 | female | heart left ventricle tissue |
| Heart | ENCFF911RNV | 1 | 0 | 100.00 | 0.00 | 59 | female | right cardiac atrium tissue |
| Colon | ENCFF886FZQ | 1 | 0 | 100.00 | 0.00 | 59 | female | left colon tissue |
| Colon | ENCFF171AQO | 11 | 9 | 18.18 | 81.82 | 40 | male | mucosa of descending colon tissue |

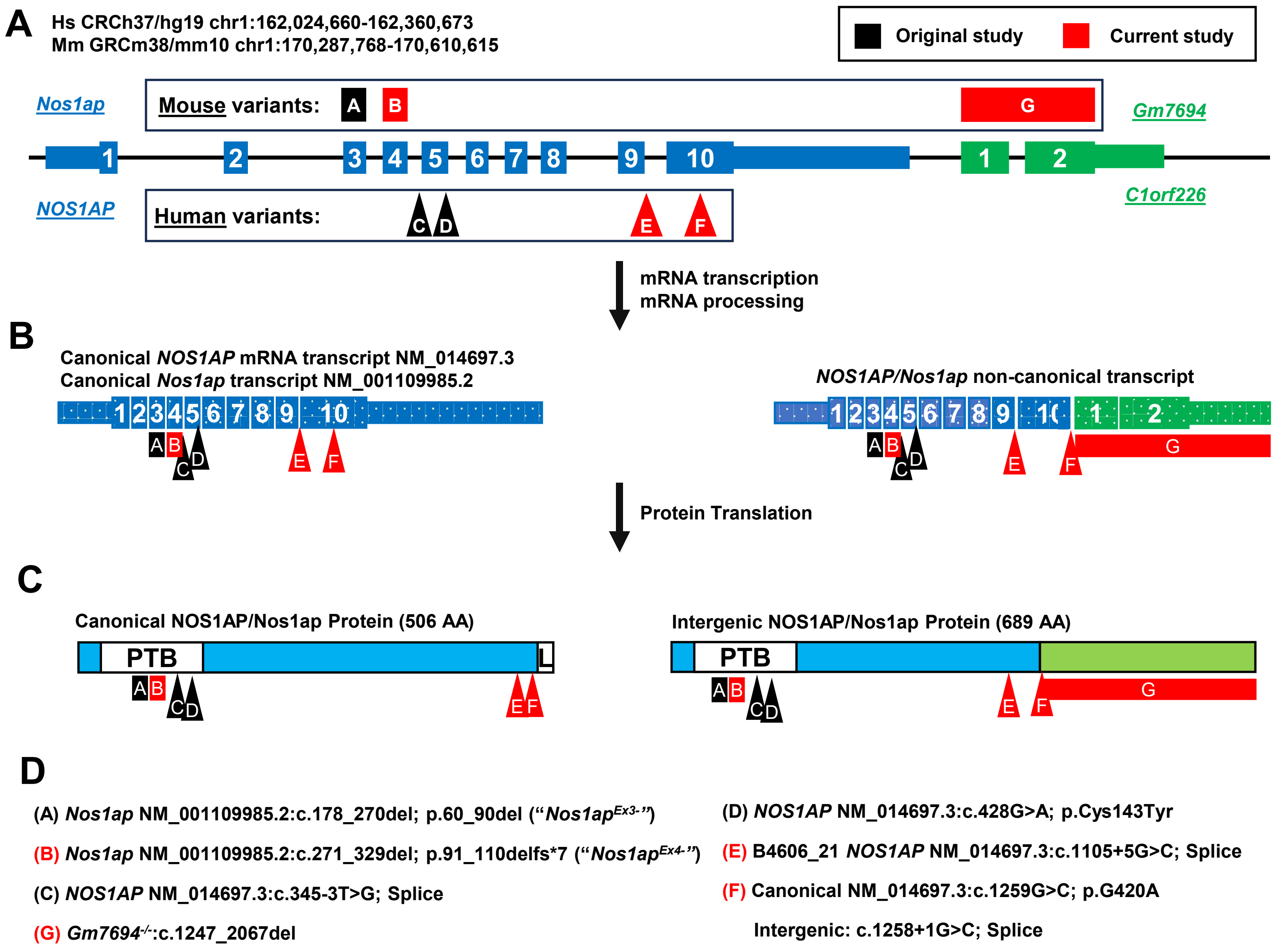


### Supplementary Figure S1

**Supplementary Figure 1. Model of *NOS1AP/Nos1ap* locus in humans and mice, resulting transcriptional and translational products, and impact of human/murine NS-associated variants.**

(A) Cartoon is shown depicting the *NOS1AP/Nos1ap* genomic locus in humans/mice. Exons are indicated that contribute to the canonical and a non-canonical transcript. The latter results from intergenic splicing that joins 164 nucleotides from exon 10 of *NOS1AP/Nos1ap* to exons 1 and 2 of *c1orf226/Gm7694*. Human and murine variants associated with nephrotic syndrome are shown as arrowheads (single nucleotide variants) or rectangles (large deletions). Shading distinguishes previously published variants (**black**) from novel variants reported in this article (**red**). Numbering is assigned to each variant as described in (D).

(B) The mRNA transcriptional products are shown with the canonical transcript NCBI accession number. Variants are similarly shown.

(C) The resulting protein products from translation are shown containing PTB domain and, in case of canonical protein, NOS1 PDZ binding domain (L). Variants are similarly shown.

(D) Human and murine variants are listed that were previously published (**black**) or are novel variants reported in this article (**red**).

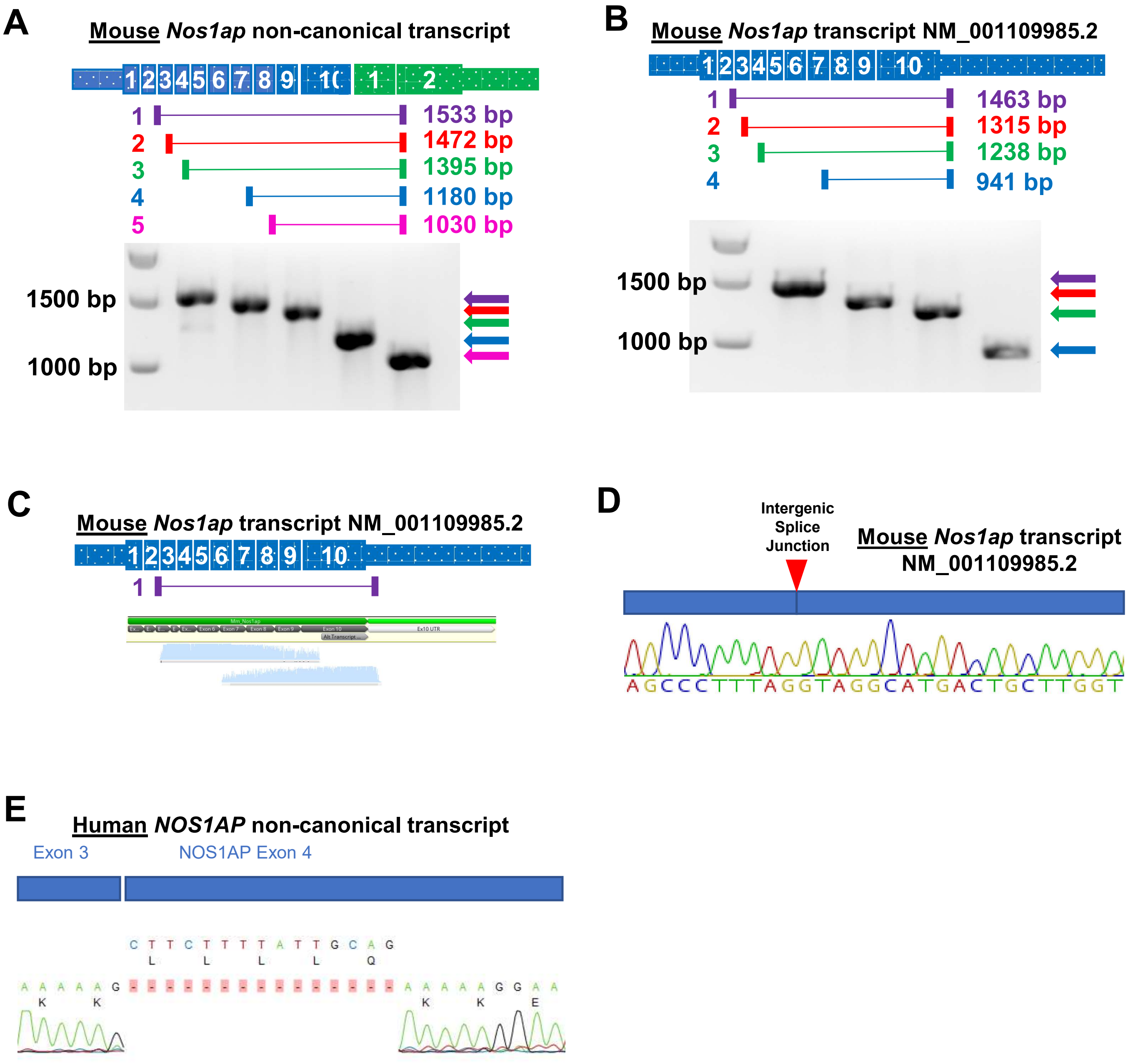


Supplementary Figure S2

**Supplementary Figure S2. Detection of alternative *NOS1AP/Nos1ap* transcripts in mice and human kidney by RT-PCR.**

(A) Cartoon is shown depicting the murine *Nos1ap* intergenic splice product. Exons are indicated that arise from the canonical *Nos1ap* (**blue**) or adjacent *Gm7694* (**green**) loci. The non-canonical transcript results from intergenic splicing that joins the first 153 nucleotides from exon 10 of *Nos1ap* to exons 1 and 2 of *Gm7694*. An amplicon spanning from early *Nos1ap* into exon 2 of *Gm7694* which was identified by RT-PCR in adult mouse kidney total RNA is depicted below the transcript.

(B) Cartoon is shown depicting the murine *Nos1ap* canonical splice product. Exons are indicated that arise from the canonical *Nos1ap* (**blue**) locus. Amplicons 1-4 are depicted below the transcript, which were identified by RT-PCR in from adult mouse kidney total RNA.

(C) Sanger sequence reads of product 1 in (A) was aligned to the canonical *Nos1ap* transcript, demonstrating a contiguous transcript from early exons into canonical 3' UTR.

(D) Sanger sequencing plot resulting from amplicon 1 in (A, B) is shown demonstrating mRNA sequence that extends across the intergenic splice junction, indicating this reflects the canonical sequence.

(E) Sanger sequencing plot resulting from the 1943 bp amplicon in (D) is shown demonstrating that within the fusion transcript, exon 4 is exclusively detected without a sequence, coding for amino acids LLLLQ, that is optional in the canonical *NOS1AP* transcript.

**A**

##### 5 Basepair Sliding Window Method

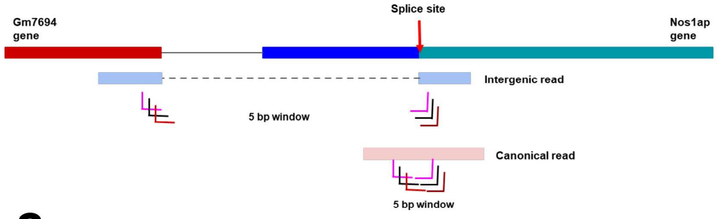


**B**

##### 10 Basepair Fixed Window Method

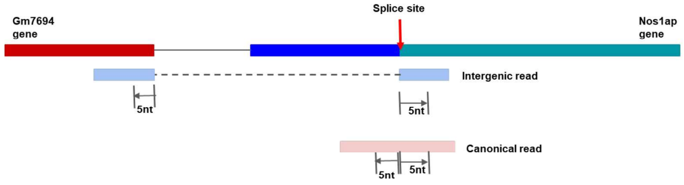


**C**

##### Summary Data for GSE225622 Dataset

| Samples and Datasets Aggregated (Xiong *Dev Cell* 2023) | Aggregate Sequence Depth (million reads) | Canonical Reads (CR) (5 BP Sliding Window) | Intergenic Reads (IR) (5 BP Sliding Window) | Ratio *Nos1ap* IR / Total Reads (5 BP Sliding Window) | Canonical Reads (CR) (10 BP Fixed Window) | Intergenic Reads (IR) (10 BP Fixed Window) | Ratio *Nos1ap* IR / Total Reads (10 BP Fixed Window) |
|---|---|---|---|---|---|---|---|
| 8 Newborn Mice 8 datasets | 258.5 | 12 | 4 | 0.25 | 8 | 4 | 0.33 |
| 10 2-week-old Mice 30 datasets | 365.9 | 11 | 4 | 0.27 | 7 | 3 | 0.30 |
| 10 4-week-old Mice 30 datasets | 357.3 | 2 | 4 | 0.67 | 2 | 4 | 0.67 |
| 10 8-week-old Mice 30 datasets | 359.7 | 2 | 8 | 0.80 | 2 | 8 | 0.80 |
| 9 79-week-old Mice 9 datasets | 271.5 | 2 | 4 | 0.67 | 2 | 3 | 0.60 |

**D**

##### Summary Data for GSE145053 Dataset

| Samples and Datasets Aggregated (Conway JASN 2020) | Aggregate Sequence Depth (million reads) | Canonical Reads (CR) | Intergenic Reads (IR) | Ratio *Nos1ap* IR / Total Reads |
|---|---|---|---|---|
| 1 8-week-old mouse 3 datasets | 34.8 | 0 | 14 | 1 |
| 1 8-week-old mouse 3 datasets | 35.5 | 0 | 15 | 1 |
| 1 8-week-old mouse 3 datasets | 38.1 | 3 | 21 | 0.875 |
| 1 8-week-old mouse 3 datasets | 36.1 | 0 | 13 | 1 |

**E**

##### Percentage of Reads in Dataset GSE145053 (5 BP Sliding Window)

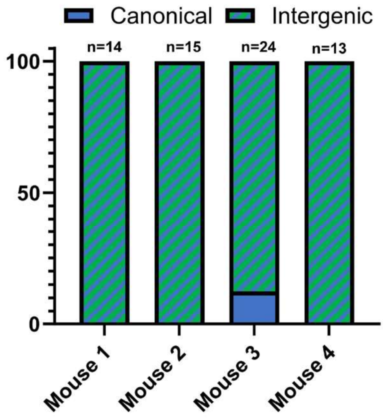


#### Supplementary Figure S3

**Supplementary Figure S3. Detection of RNA transcript reads of canonical and intergenic *Nos1ap* transcripts from mouse kidney.**

(A) Cartoon describing the criteria for including reads spanning the canonical or intergenic *Nos1ap* transcripts. Using the sliding 5 Basepair Sliding Window, reads crossed the intergenic splice junction (chr1: chr1:170318737-170318738) and span a minimum of 5 nucleotides on either side of the transcript.

(B) As in (A), criteria for read calling in mouse RNA sequencing data is shown. For the 10 Basepair Fixed Window, canonical reads are counted if they span minimum of 10 nucleotides from region chr1:170318733-170318742 on exon 10 of the *Nos1ap* gene. Intergenic reads are counted if they span minimum of 5 nucleotides in region chr1:170302834-170302838 on exon 1 of *Gm7694* AND 5 nucleotides in region chr1:170318738-170318342 on exon 10 of *Nos1ap.*

(C) A table is shown displaying the ratio of canonical and intergenic reads in RNA sequencing data from 0, 2, 4, 8, and 79 week-old C57BL/6 mice are shown using a 5 bp sliding window approach or using a 10 bp fixed window approach for dataset GSE225622.

(D) A table is shown displaying the ratio of canonical and intergenic reads in RNA sequencing data from four 8-week-old C57BL/6J mice are shown using a 5 bp sliding window approach for dataset GSE145053.

(E) The ratio of canonical and intergenic reads in RNA sequencing data from four 8 week-old mice (dataset GSE145053) are shown using a 5 bp sliding window approach. Reads (n) are shown above box plot and derived from (D).

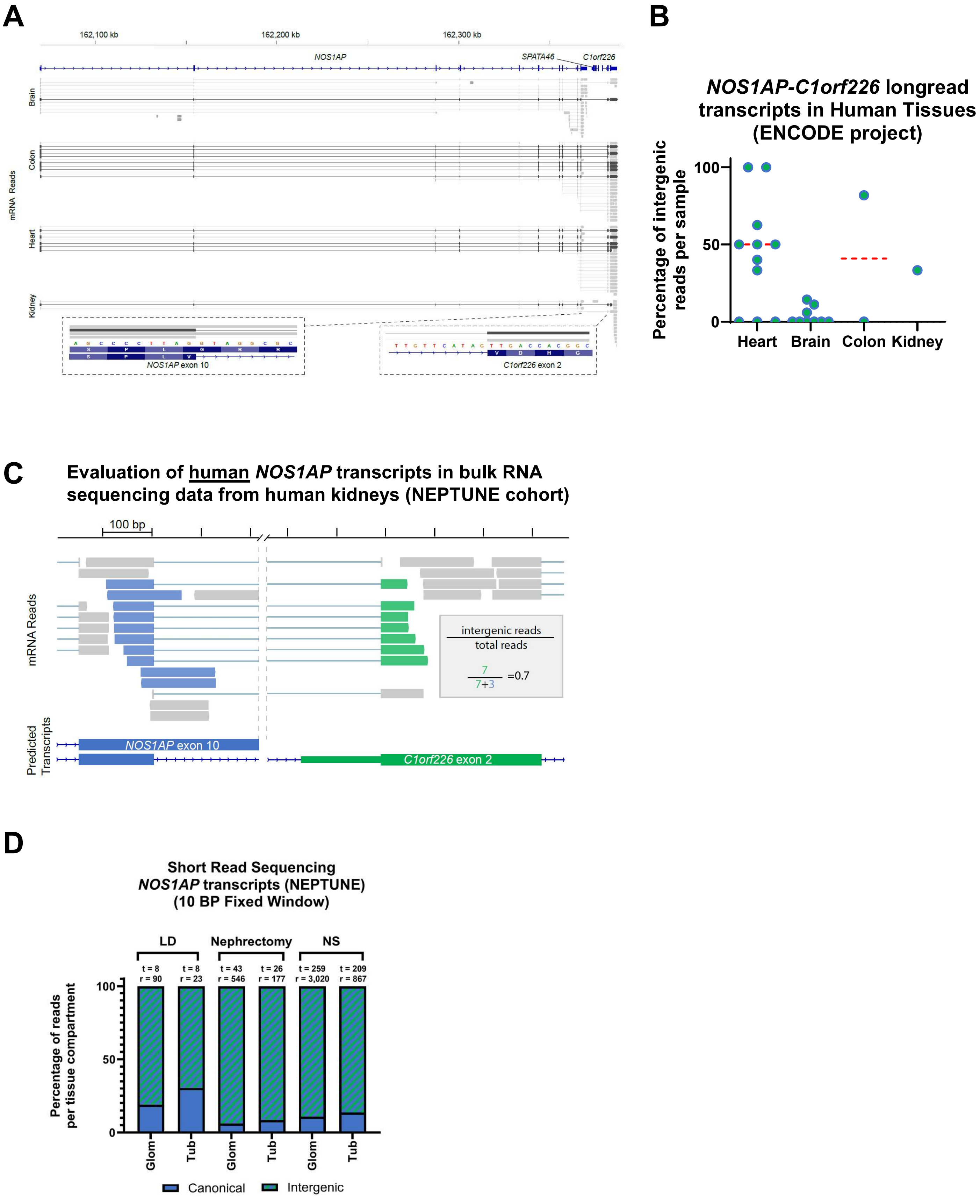


### Supplementary Figure S4

**Supplementary Figure S4. Detection of RNA transcript reads of canonical and intergenic *Nos1ap* transcripts from human tissues including kidney.**

(A) Example of long read sequencing data from human tissue samples is shown aligning to the *NOS1AP-C1orf226* locus. Single samples from heart, brain, colon and kidney are shown. Dark grey reads represent intergenic reads beginning within or before *NOS1AP* exon 3 and crossing into *C1orf226.* Light grey reads represent canonical reads beginning within or before *NOS1AP* exon 3 and ending after the intergenic splice junction in *NOS1AP* exon 10.

(B) The dot plot displays percentage of intergenic *NOS1AP* transcripts for each sample in ENCODE long read RNA sequencing data. Samples are grouped by tissue type.

(C) A cartoon is shown to depict how reads with a fixed 10 base pair window around the intergenic splice junction in *Nos1ap* were quantified as intergenic (blue-green) or canonical (blue only) and quantified as a ratio.

(D) Box plot plot displays percentage of *NOS1AP* transcripts reflecting canonical versus intergenic splice products for each tissue compartment in NEPTUNE cohort short read RNA sequencing data. Tissues (t) and reads (r) are noted above the box plot.

### Supplementary Figure 5

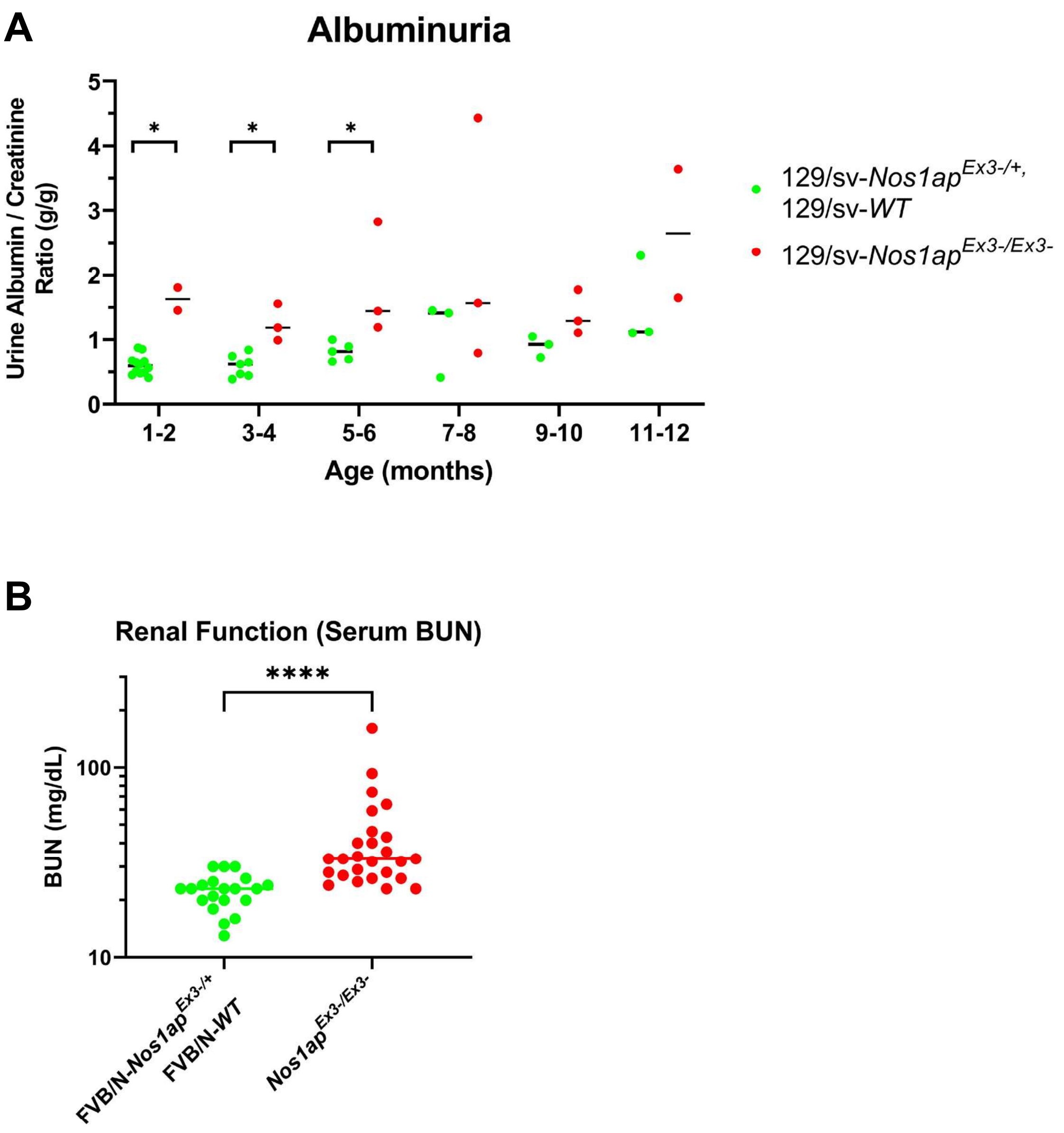

**Supplementary Figure 5. Assessment of proteinuric disease in *Nos1ap*$^{Ex3-/Ex3-}$ mice on distinct genetic backgrounds.**

(A) *Nos1ap*$^{Ex3-/Ex3-}$ mice on 129/sv background exhibit elevated urine albumin-to-creatinine ratios (ACR) compared to control animals. *Nos1ap*$^{Ex3-/Ex3-}$ group consists of 4 mice. Control mice consisted of 14 *Nos1ap*$^{Ex3-/+}$ heterozygous mice as well as 1 wildtype littermate. Graph shows dot plots with median bars for each genotype and time-point. Red, homozygous animals; Green, wild type and heterozygous littermate controls. Mann-Whitney test performed to compare groups at each timepoint (*$p<0.05$).

(B) Serum BUN levels were quantified in homozygotes (red) or littermate controls (green) from 3-6 months of life. Mann-Whitney test performed to compare groups (*$p<0.0001$).

### Supplementary Figure 6

A

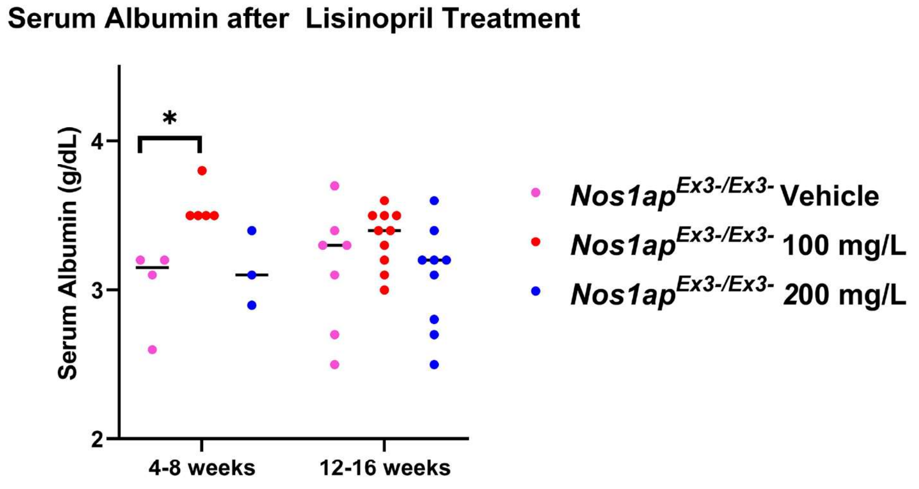


B

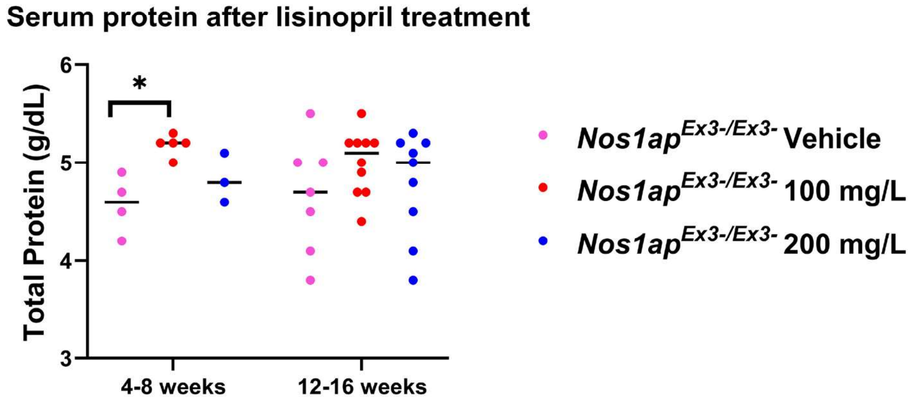


C

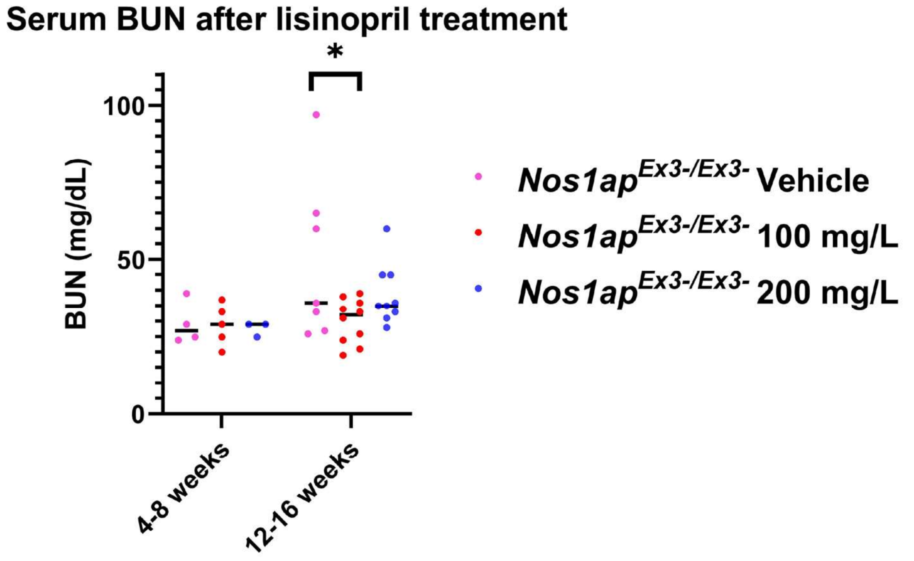

**Supplementary Figure 6. Anti-proteinuric treatment effects on serum albumin, protein, and blood urea nitrogen (BUN) levels in in FVB/N-*Nos1ap*$^{Ex3-/Ex3-}$ mice.**

FVB/N-*Nos1ap*$^{Ex3-/Ex3-}$ mice with comparable baseline ACRs received drinking water with either vehicle (water), 100 mg/L lisinopril, or 200 mg/L lisinopril starting at 6 weeks of age. Serum was collected at 4-8 weeks and 12-16 weeks of treatment. (A) Serum albumin, (B) serum total protein, and (C) Blood urea nitrogen (BUN) were quantified using the VetScan VS2 system and "Comprehensive Diagnostic Profile" rotors. Animals treated with 100 mg/L lisinopril showed significantly increased levels of serum albumin and total protein relative to vehicle treated mice mice after 4-8 weeks of treatment and significantly decreased BUN levels after 12-16 weeks of treatment. No significant differences were noted between vehicle and 200 mg/L lisinopril treated groups at these timepoints. Pink, Vehicle control; Red, 100 mg/L lisinopril group; Blue, 200 mg/L lisinopril group. Student t-test performed to compare groups (*p<0.05).

### Supplementary Figure 7

**A**

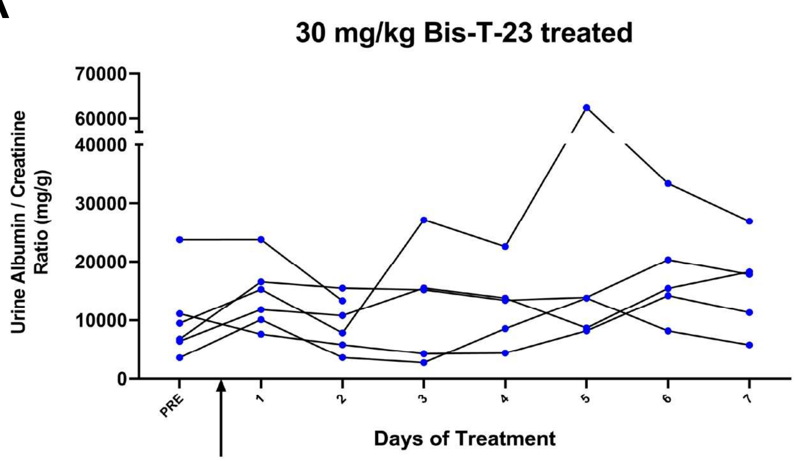


**B**

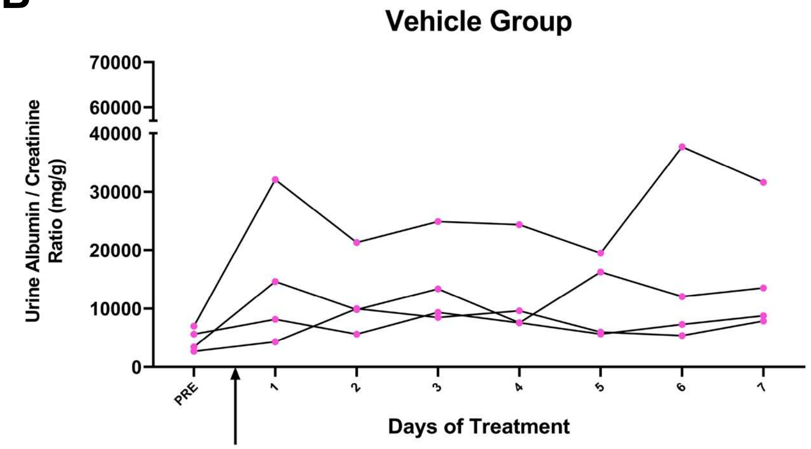


**C**

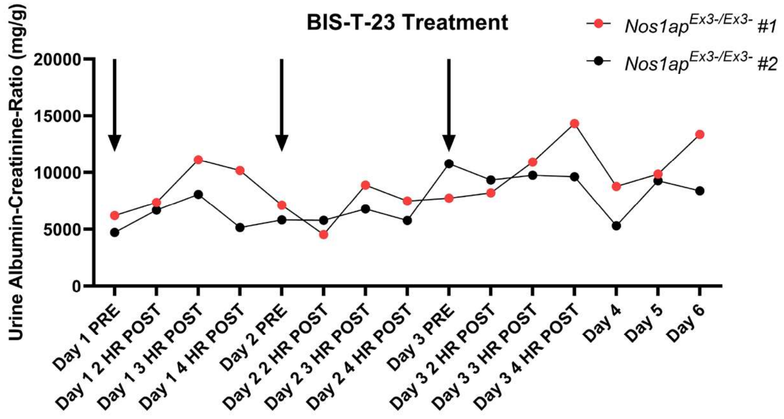

**Supplementary Figure 7. Effect of dynamin activating compound Bis-T-23 effects on albuminuria in FVB/N-*Nos1ap*$^{Ex3-/Ex3-}$ mice.**

(A) FVB/N-*Nos1ap*$^{Ex3-/Ex3-}$ mice at 4 weeks of life with comparable baseline albumin-to-creatinine ratios (ACRs) were administered intraperitoneal 30 mg/kg Bis-T-23 (in 20% DMSO/80% PBS) as marked by arrow. Urine samples prior to administration and every day following for 7 days were collected. Albumin-to-creatine ratios were measured and were not reduced upon Bis-T-23 injection.

(B) FVB/N-*Nos1ap*$^{Ex3-/Ex3-}$ mice at 4 weeks of life with comparable baseline albumin-to-creatinine ratios were administered the vehicle for (A) (20% DMSO/80% PBS) as marked by arrow. Urine was collected in parallel with samples in (A). ACR were not reduced with vehicle administration.

(C) Two FVB/N-*Nos1ap*$^{Ex3-/Ex3-}$ mice at 4 weeks of life with comparable baseline albumin-to-creatinine ratios (ACRs) were injected with 20 mg/kg intraperitoneal Bis-T-23 on three consecutive days (see arrows). Urine was collected at 2, 3, and 4 hours after injection on days 1-3 and subsequently on days 4, 5, and 6. No reduction in ACR were detected.
